# Phase II Clinical trial for Evaluation of BCG as potential therapy for COVID-19

**DOI:** 10.1101/2020.10.28.20221630

**Authors:** Usha Padmanabhan, Sanjay Mukherjee, Rohidas Borse, Sameer Joshi, Rajesh Deshmukh

**Author notes:** Both authors must be considered as first authors who contributed equally to the work (Usha Padmanabhan, Sanjay Mukherjee). Corresponding author: Dr. Usha Padmanabhan, Address: Biochemistry department, Haffkine Institute for Training, Research & Testing, Acharya, Donde Marg, Parel, Mumbai 400 012., Contact.

## Abstract

Bacillus Calmette−Guérin (BCG) is widely used in national vaccination programs worldwide. It is accepted that BCG alleviates both pathogen and allergy induced respiratory diseases that could also include Covid-19. To investigate this possibility, we randomly assigned 60 Covid-19 patients, after admission to the hospital with pneumonia and requirement for oxygen therapy in a 1:1 ratio to receive either a single adult dose of intradermal BCG or normal saline with concomitant standard of care (SoC) medications. Primary endpoints were favorable prognosis of Covid-19 as deduced from resolution of pneumonia, viremia and secondary outcome were enumeration of ICU admissions, duration thereof and mortalities.

**Results:** Both primary and secondary endpoints were significantly improved in the BCG+SoC group. This could be seen from reduction in oxygen requirement due to Covid-19 associated pneumonia decreasing from day 3-4, improved radiological resolution from day 7-15. There were a total of 6 (10%) adverse events in the study of which 2 deaths and 4 ICU admissions were in SoC group (1 ICU admission culminated in death of the subject) and in contrast only 1 ICU admission in the BCG+SoC group. While there was an increase in Covid-19 specific IgG levels in the BCG+SoC group, there was no evidence of BCG induced cytokine storm in this group. Four patients showed localized inflammatory response at the injection site in the BCG+SoC group.

**Conclusions:** BCG+SoC administration resulted in a significantly higher percentage of patients with favorable outcomes than did SoC. A third of the patients were naïve for childhood BCG vaccination. This mimicked elderly patients in countries with no universal vaccination policy for BCG. No BCG related adversity was seen in this group. The study shows that BCG is a safe, cost-effective treatment that can be introduced as a standard of care in patients with moderate Covid-19 that can reduce requirement of oxygen supplemented beds and disease burden in low resource countries, with additional long-term benefits of reducing risk for tuberculosis.

## Introduction

The novel coronavirus nCoV-19 or 2019-nCoV (severe acute respiratory coronavirus 2 or SARS-CoV-2), has affected more than 10 million people worldwide, resulting in significant mortality and is responsible for the global pandemic COVID-19. The clinical course is variable of COVID-19 is variable with older age and the presence of comorbidities increasing the risk of more severe disease [1-3].

*BacilleCalmette-Guérin* (BCG) is a live attenuated strain of the bovine tuberculosis bacillus, *Mycobacterium bovis*, that is the only WHO recommended vaccine against tuberculosis till date shown to have non-specific protective effects against allergenic and pathogenic respiratory disorders [4-5]. Miller et al, Hegarty and Dayal et al., 2020 [6-8] show a negative correlation between BCG immunization status of a country and mortalities due to COVID-19. Epidemiological data presented therein that suggested that BCG could be effective against nCoV-19 or SARS-CoV-2. It was the purpose of this study to investigate a direct link between BCG intervention in moderate Covid-19 patients with primary endpoints of reduction in oxygen dependency, faster resolution of COVID-19 symptoms, an evaluation of viremia and favorable outcome for COVID-19.

## Methods

### Ethics and Permissions

The trial was an investigator-initiated, individually randomised, placebo-controlled, double-blind interventional phase II trial to assess the effectiveness and safety of intradermal BCG in adults (aged 18-60 years) admitted to hospital with COVID-19. Ethical approval was obtained from the Institutional Ethics Committee (IEC) of the BJ Medical College & Sassoon Hospital, Pune. The trial was also approved by the Drug Controller General of India (DCGI). The trial was registered with Clinical Trial Registry of India no. **CTRI/2020/05/025013**. (CTRI is a Primary Register of the International Clinical Trials Registry Platform (ICTRP, http://www.who.int/ictrp/search/en/). The trial was done in accordance with the principles of the Declaration of Helsinki and the International Conference on Harmonization–Good Clinical Practice guidelines. The final approved protocol is available along with this article (Supplementary Appendix). Written informed and videographic consent was obtained from all patients. The authors vouch for the accuracy, completeness of data and the reporting of adverse events and for the fidelity of the trial to the protocol.

### Study design

#### Subject and randomization

Eligible Subjects (n=60) were adults; men and non-pregnant women (aged 18-60 years) with COVID-19 admitted to hospital with laboratory-confirmed SARS-CoV-2 infection (as pre RT-PCR), with an interval from symptom onset to enrolment of 12 days or less, oxygen saturation of 94% or less on room air and radiologically confirmed pneumonia. Exclusion criteria included any co-morbidities, immunodeficiency disorders, pregnant and lactating women. Subjects were screened and enrolled in multiples of two, so that subjects could be randomized parallely into both arms. Double blind randomization was achieved by on-site physician choosing identically loaded syringes with saline or reconstituted BCG (see Consort Flow Diagram Fig S1, Fig S2 and Protocol in Supp Appendix).

#### BCG and Standard of Care (SoC)

Patients were randomly assigned in a 1:1 ratio to the BCG arm (single intradermal adult dose of 0.1 ml of 2.0 - 8.0 × 10^6^ c.f.u. on enrollment day assigned as day 0, hereinafter) or SoC arm who received the same volume of normal saline. Concomitant use of SoC was approved by DCGI at the time of granting permission for the Clinical trial and SoC medications were decided by the treating on-site physician and as per the guidelines issued by the Indian Council for Medical Research (ICMR) from time to time (see Supp Appendix).

### Study Assessments

The study assessments included daily multiple measurements of SpO2, pulse rate and other vital sign measurements, periodic clinical laboratory testing (CBC, serum ferritin, CPK-MB, biochemical parameters) and Chest X rays. Adverse events such as ICU admission, non-invasive ventilation, intubation, death leading to discontinuation of treatment were reported to the IEC and DCGI initially within 24 hours and then reported in detail by 14 days. Blood sampling and nasopharyngeal swabs in VTM for evaluating viremia were collected on days 0, 7 and 15. Clinical data were recorded on paper case record forms and then entered into an electronic database.

### Viral load assessment

Nasopharyngeal swabs were collected in VTM for viral RNA detection and quantification. VTMs were stored at −80°C until they were thawed for analyses. The viral load was estimated using RT-qPCR as per details in the Supplementary Appendix.

### Cytokine analyses

Serum levels of IL-6, TNF-alpha, IFN-gamma, IgG and IgM (nCov-19 specific) were evaluated from blood samples of days 0, 7 and 15 using ELISA kit from Krishgen Biosciences and Raybiotech, as per manufacturer’s instructions.

### Statistical analyses

In general, for analyses between the two major groups in the study BCG+SoC and SoC, median values of data for each group were considered and Mann Whitney or Wilcoxon Signed Rank analyses as stated was employed. Where there were more than 2 groups, Kruskal Wallis analyses was performed. Patients who had not received BCG vaccination (which is part of the Universal vaccination program in India) as neonates, hereinafter referred to as naïve patients, were post-hoc analyzed as a separate subgroup. Statistical differences were considered significant when p<0.05. All probabilities are two-tailed. All statistical tests and graphs were performed by GraphPad Prism 5.0 (GraphPad software, San Diego, CA).

## Results

### PATIENTS

Table 1 outlines the demographic and clinical characteristics of the total of 60 patients who were enrolled in the study, 36 were males (60%) and 24 (40%) were females. Age analyses revealed that patients randomized to receive BCG+SoC (median age = 49 yrs) were older than those who received SoC (median age = 41.5 yrs, p = 0.076, Mann Whitney analyses). About 40% patients were obese/overweight (26/60). Of these 20% (12/30) were obese (9 on BCG+SoC, 3 on SoC) and (12/30) 20% were overweight (4 on BCG+SoC, 8 on SoC).

**Table 1:** Primary analyses of subjects in the trial.

| **Description** | **Total** | **SoC** | **BCG** |
| --- | --- | --- | --- |
| **Total (N)** | 60 | 30 | 30 |
| **Males** | 36 (60%) | 16 | 20 |
| **Females** | 24 (40%) | 14 | 10 |
| **Median Age (yrs)** |  | 41.5 (21-60) | 49 (26-60) |
| **Overweight (BMI 25-30)** | 12 (20%) | 9 | 3 |
| **Obesity (BMI > 30)** | 14 (23%) | 3 | 11 |
| **No. of ICU admission for monitoring** | 5 (8.3%) | 4 | 1 |
| **Death** | 2 (3.33%) | 2 | 0 |
| **Withdrawal** | 2 (3.33%) | 1 | 1 |
| **Delayed Hypersensitivity Skin response to BCG** | 4 (6.67%) | 0 | 4 |
| **Secondary Herpes infection** | 2 | 1 | 1 |
| **Hypothyroidism** | 5 (8.3%) | 3 | 2 |
| **Standard of care** | 60 | 30 | 30 |
| **Remdesivir** | 5 (6.67%) | 4 | 1 |

SoC medications were as per the national guidelines and were not contraindicated with concomitant use of BCG (see Fig S3). Remdesivir was administered to four (4/30, 13.33%) patients on the SoC arm as an emergency care option as specified by ICMR [9]. Four (4/30, 13.33%) patients in the BCG+SoC group showed local inflammatory response to BCG (see Fig S5A).

### PRIMARY OUTCOME

#### Resolution of Covid-19 associated hypoxia

The SpO2/FiO2 data of subjects in the BCG+SoC and SoC groups were compared (Fig 1A,B). The BCG+SoC group had a significantly better SpO2/FiO2 values compared to the SoC group (p = 0.0345, Mann Whitney analyses). Linear regression analyses of the data (Table S1) indicates that rate of increase in SpO_2_/FiO_2_ levels per day for BCG+SoC = 13.16±1.22; for SoC = 6.55±1.94 (p = 0.0026 that rates are different), indicating that BCG+SoC had significantly faster resolution of Covid-1 associated hypoxia.

**Figure 1:**
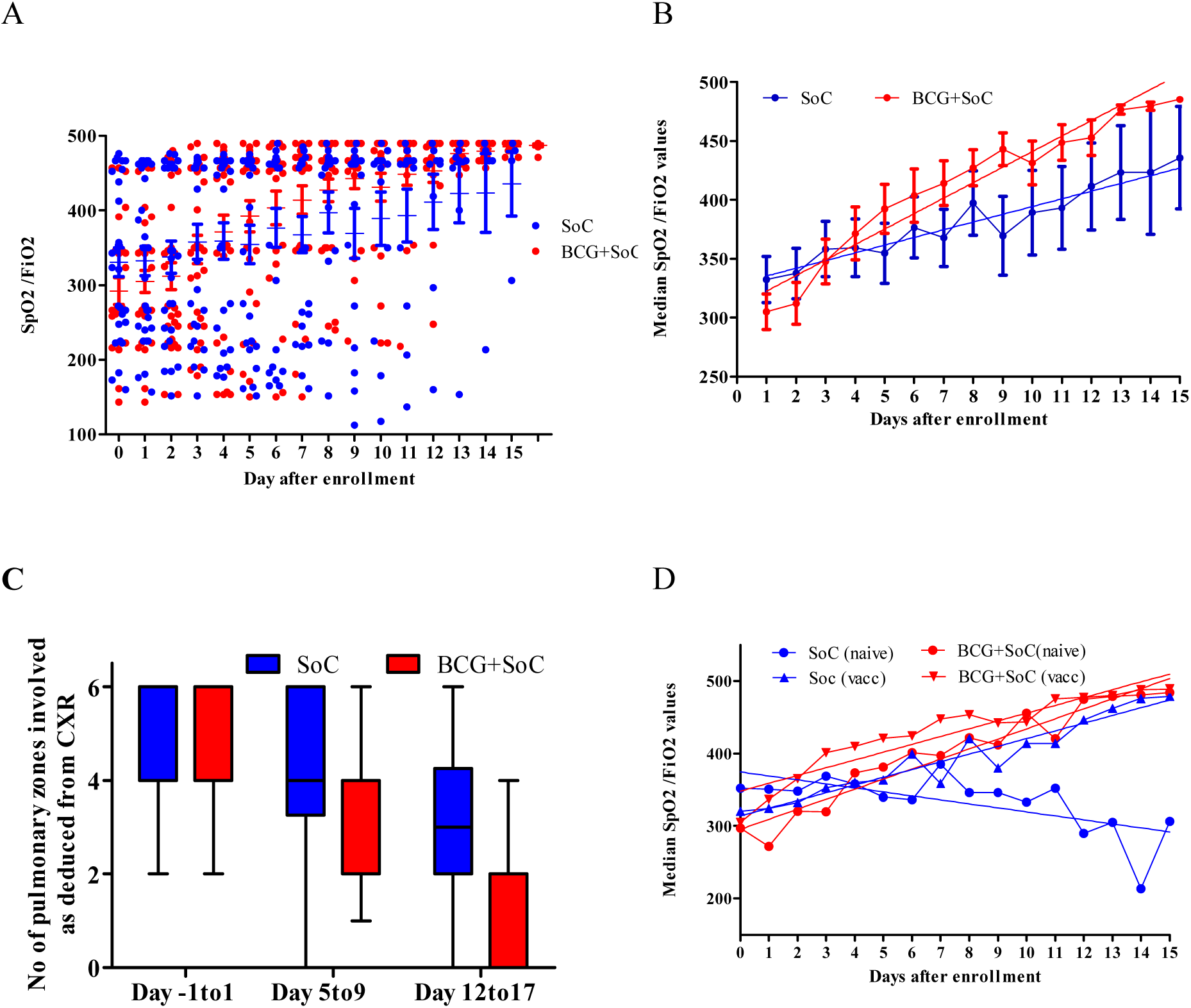
Effect of BCG+SoC administration on oxygen saturation. **(A)** Each point indicates SpO2/FiO2 calculated from measured SpO2 for each subject from enrollment on that corresponding day. Since SpO2 was measured for each subject several times in a day, each point is an average of all SpO2 measurements for that subject on that day. All subjects on BCG arm were administered WHO recommended adult dose of BCG (0.1 ml of 2-8 ×10^6^ CFU,) on Day 0 of enrollment. **(B)** Each point represents median of 30 data points on the corresponding day. Vertical bars represent min-max ranges of medians calculated for that corresponding day. The black lines represent the linear regression lines fitted to the data. **(C) Effect of BCG administration on Covid-19 associated pneumonia as deduced from CXR of patients**. Each bar represents number of zones involved as deduced from the chest X rays of the patients on the corresponding day. Vertical ranges represent minimum-maximum ranges **(D) Effect of BCG administration in** O**xygen saturation status in naïve and vaccinated patients**. Each point indicates median of SpO2/FiO2 data points [SoC (vacc) group, 21 pts, blue triangles; BCG+SoC (vacc) group, 20 pts, red circles; SoC (naïve) group, 9 pts, blue circles and BCG+SOC (naïve) group, 10 pts, red triangles] as calculated from measured SpO2 for each subject on that corresponding day. Vertical bars represent min-max ranges of medians calculated for that corresponding day. The black lines represent the linear regression lines fitted to the data.

#### Radiological Resolution of Covid-19 pneumonia

As per the Fleischner Society consensus statement, the typical CT findings of COVID-19 are bilateral and peripheral predominant ground glass opacities [10-11]. CXR images were evaluated for the presence and distribution of the following abnormalities hazy increased lung opacity in number of zones involved, hilar abnormalities, cardiomegaly and pleural effusion. As shown in Fig 1C, over a 15 day period, subjects in the BCG+SoC arm showed significant radiological improvement over the SoC arm in concurrence with the improvement seen in oxygen saturation.

#### BCG Naïve Subjects & Covid-19 associated hypoxia

About a third (19/60, 30%) of the subjects in the study were naïve and had not received BCG vaccination as neonates. Of these 9 (5 males and 4 females) were randomized to the SoC arm and 10 (4 males and 6 females) were in the BCG+SoC arm. Linear regression analyses of the data (Fig 1D and Table S2) indicate that rate of increase in SpO_2_/FiO_2_ levels per day for BCG+SoC(naïve) = 13.87±1.05; for SoC(naïve) = −5.51±1.69; BCG+SoC(vacc) = 10.68±0.85; for SoC(vacc) = 10.76±1.07 (p < 0.001, Kruskal wallis test).

Analyses of the age of these subjects indicated that age of naïve subjects [median age = 54.5 yrs (21-60 yrs)] in the study was significantly higher (p = 0.0033, MannWhitney test) than that of vaccinated subjects [median age = 44 yrs (26-60 yrs)]. Amongst the naïve subjects, there was no significant difference in ages of subjects in the BCG+SoC arm [median age = 55.5 yrs (21-60 yrs)] and the SoC arm [median age = 55 yrs (21-60 yrs), p = 0.64, MannWhitney test] (Fig S3B&C).

#### Viremia

We checked whether administration of BCG would have an effect on the viral load in patients by estimating the C_T_ values (Supp Fig S6). Supp Fig S6 A&B show the amplification curves for subjects enrolled from 6th May - 15th Jun 2020 and from 15th Jun - 5th Aug 2020, respectively. The melt curves in Fig. S6C show a single peak indicating specificity of primers. Shown in Fig. 2A are the C_T_ values of subjects enrolled from 6th May −15th Jun 2020 on (a) SoC arm SoC-0 & SoC-7 (days 0&7) were 20.77±3.59, 23.21±3.71 (p = 0.078, 2 tailed Wilcoxon Signed rank test) and (b) BCG+SoC arm BCG+SoC-0, BCG+SoC-7 (days 0&7) were 19.93±2.69 & 26.007±2.9, respectively (p = 0.0078, 2 tailed Wilcoxon Signed rank test). Since C_T_ values are inversely indicative of viral load, the data indicate viral loads higher on admission and lower on day 7 with significant reduction in viral load in the BCG+SoC arm. Shown in Figure 2B, are the analyses of subjects enrolled from 15th Jun - 8th Aug 2020. C_T_ values of RT-PCR As shown in Figurer 2B, the mean C_T_ values of SoC-0, SoC-7 were 26.53±1.25, 26.01±0.57 (p = 0.053, 2 tailed Wilcoxon Signed rank test) and for BCG+SoC-0, BCG+SoC-7 were 25.61±2.57, 26.84±4.9 respectively (p = 0.72, 2 tailed Wilcoxon Signed rank test) indicating of insensitivity to BCG and/or SoC treatment.

**Figure 2:**
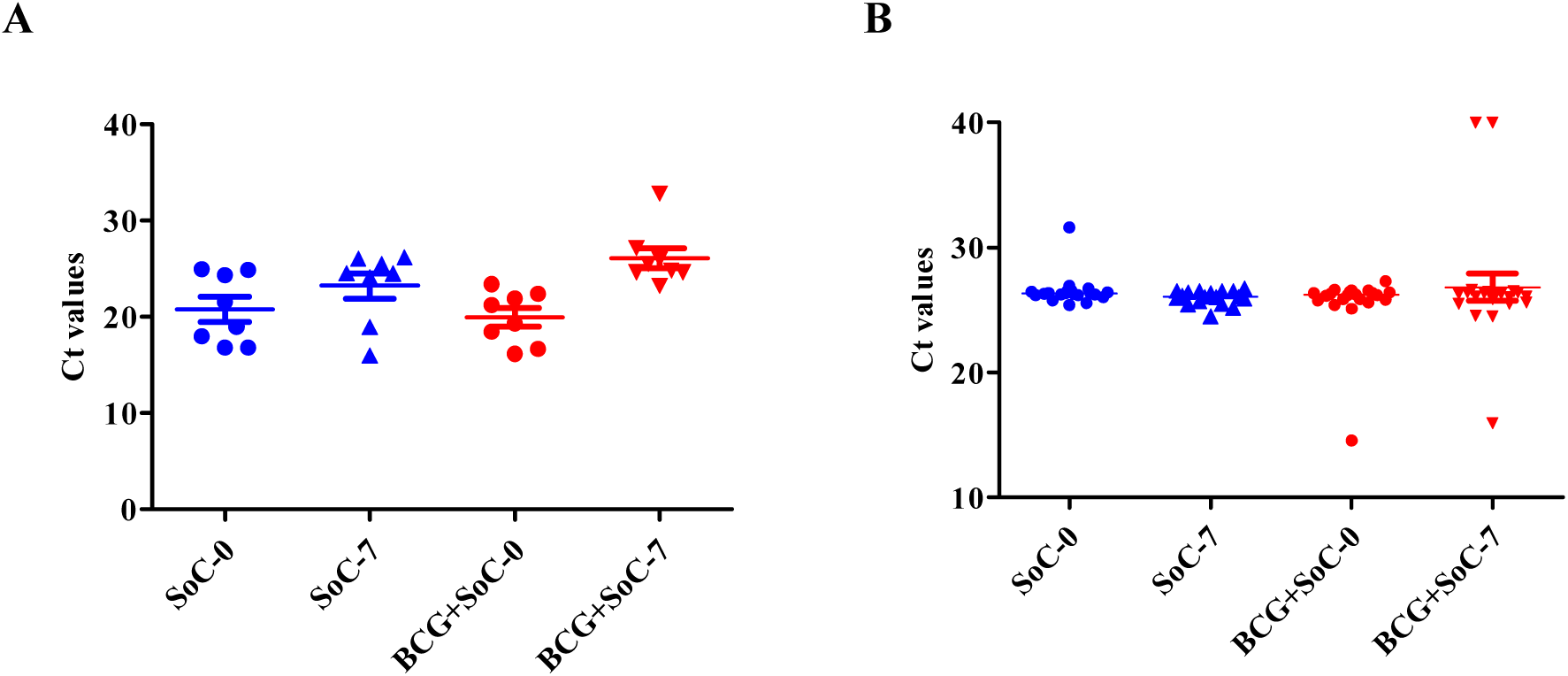
Effect of BCG on Viremia: **A)** Each dot represents C_T_ value of each of first 20 subjects who were enrolled in month of May-Jun 2020 **B)** Each dot represents C_T_ value of each of next 40 subjects who were enrolled in month of Jun-Aug 2020. Each point is an average of duplicate experiments performed. For details please see Supplementary Figure 3. SoC-0, BCG+SoC-0 and SoC-7, BCG+SoC-7 represents the C_T_ values derived from virus in VTM of nasopharyngeal swabs from patients on both arms in VTM on Day 0 and Day 7, respectively **SoC**: Standard of Care

### SECONDARY OUTCOMES

Table 1 shows that the number of ICU admissions were 5 out of which 4 patients were on the SoC arm and 1 in the BCG+SoC arm. There were 2 deaths in the SoC arm out of which one was an ICU admission which resulted in death and none on the BCG+SoC arm. Remdesevir was used on emergency basis in India as SoC [9] and was administered to 4 patients on the SoC arm and to 1 subject in the BCG+SoC arm (data of this patient not analyzed, see Fig S1).

#### Hematology

As shown in Figure 3 (A&B) overall comparison of subjects in SoC and BCG+SoC revealed significantly higher WBC count (p = 0.036), a mild increase in neutrophil count in SoC group and similar lymphocyte count in both groups. Over a 15 day period, the lymphocyte count in both groups showed an increase and the NLR (neutrophil to lymphocyte ratio) showed a larger decrease in BCG+SoC compared to SoC.

**Figure 3:**
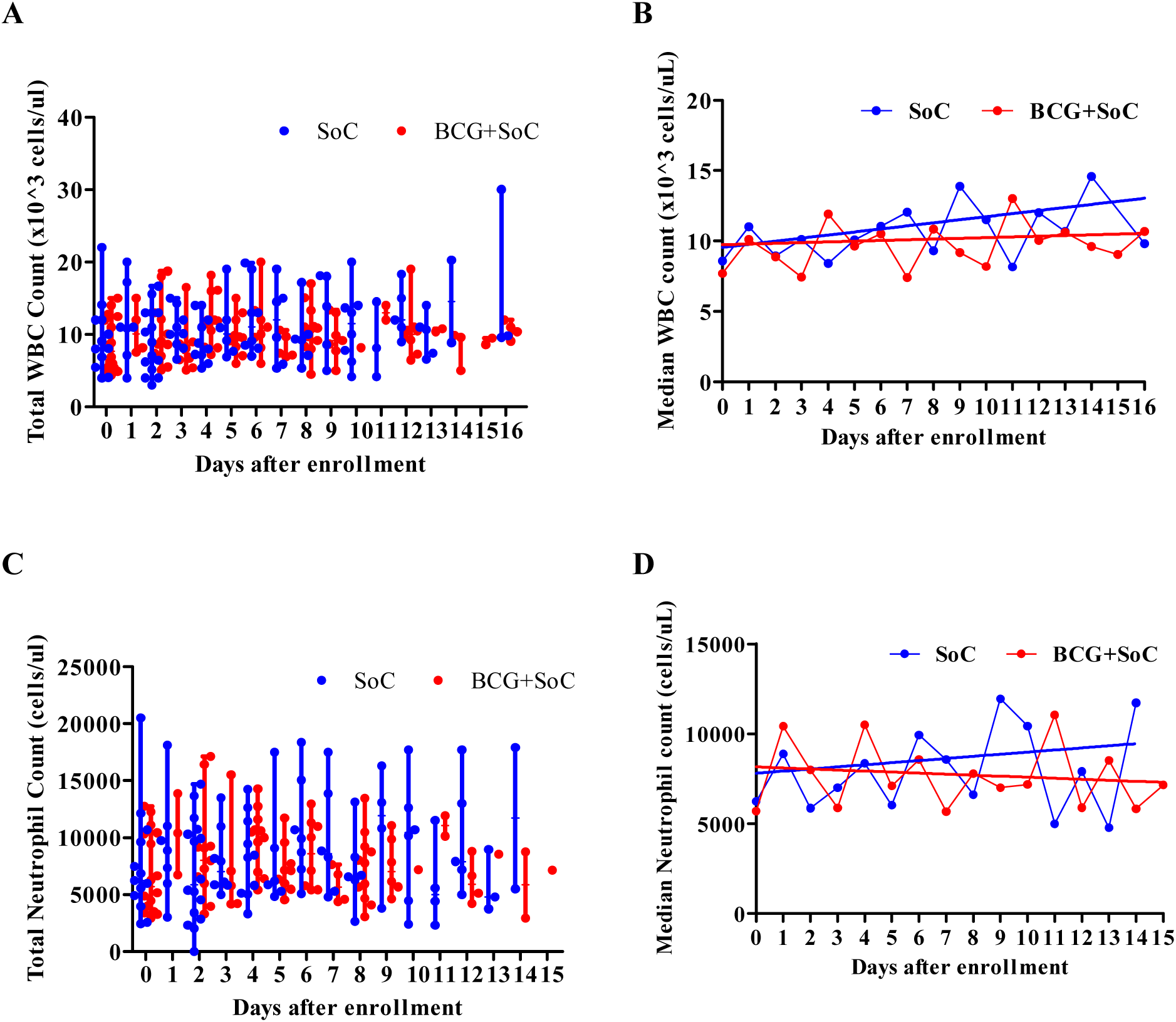
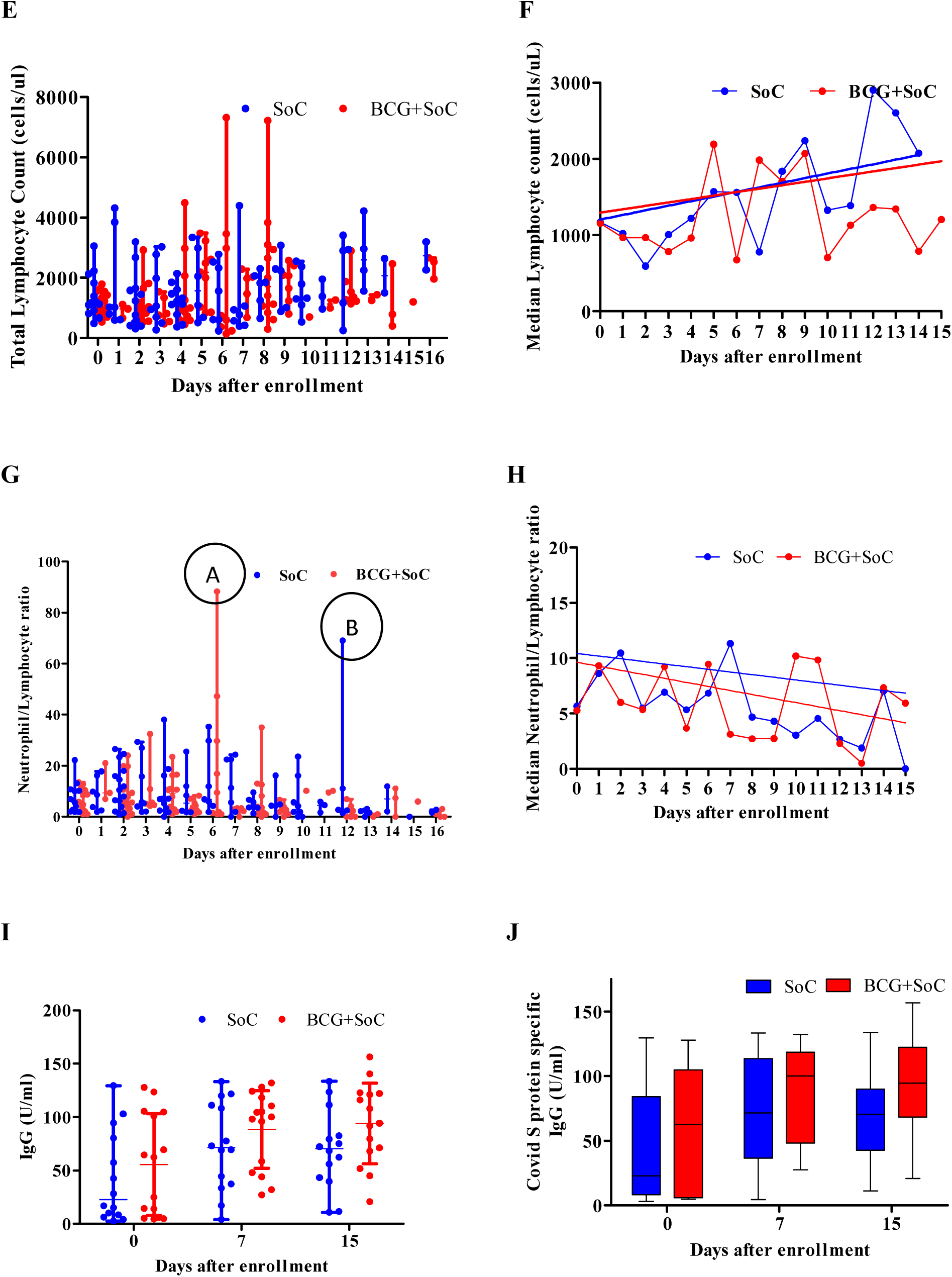
Effect of BCG on A&B: WBC count, C&D: Neutrophil count, E&F: Lymphocyte count, G&H: NLR, I &J For each panel LHS: Each point represents value of WBC, lymphocyte, neutrophil count, N/L ratio or IgG level of that subject on the corresponding day. Horizontal bars represent the median values and vertical bars represent min-max ranges of medians calculated for that corresponding day. **RHS:** Each point represents median of WBC, lymphocyte, neutrophil count, N/L ratio of 8-16 subjects on the corresponding day after enrollment. For IgG, the vertical range depicts min-max ranges, box length is 25-75%. Horizontal bars represents mean of IgG values from 30 samples on day 0, day 7 and for day 15 mean of 26 samples for SoC arm and for BCG+SoC arm mean of 28 samples on day 15. Linear regression analyses was carried out using Graphpad Prism.

NLR one of the early risk factors identified for 2019-nCoV severe illness; patients with age ≥ 50 and NLR ≥ 3.13 facilitated severe illness [12]. In our study, the NLR decreased at similar rates in both arms; median NLR was 7.19 (SoC) and 6.28 (BCG+SoC). There were 2 subjects with extremely high N/L ratio: Subject #A, N/L=88 and Subject #B, N/L= 69 (Fig 3G). In corroboration with reports that NLR is a measure of adverse prognosis of covid-19, both subjects were admitted to ICU; whereas subject #A in the BCG+SoC arm deteriorated to need NIV, subject #B on SoC deteriorated to need NIV, was subsequently intubated and later succumbed to the disease; even though subject #B had a lower N/L ratio compared to subject #A.

Comparison of WBC, neutrophil, lymphocyte count in both arms, suggests that part of the increase in WBC count in the SoC arm is due to increase in neutrophil count, which in the BCG+SoC arm is accounted by the lymphocyte count (presumably higher antibody producing B and T cells). Should this hypothesis be true, n-CoV-Sars2 specific IgM and IgG levels should be indicative of the same. Notably the total level of Covid specific IgG levels (Fig 3I & J) in the BCG+SoC arm was higher compared to SoC. However, we did not see detectable levels of n-CoV-Sars2 specific IgM (data not shown). The presence of detectable IgG, but not IgM also corroborates with timelines reported for seroconversion [13] and fairly homogenous population in this study. As our supplementary data suggests analyses of platelets, erythrocytes etc did not reveal any significant differences between the two groups (Fig S8).

### SAFETY

#### Cytokine storm and Covid-19 prognosis

Patients with severe COVID-19 show elevated inflammatory markers, including ferritin [12-14], IL-6 [15], which constitute the cytokine storm that has been associated with critical and life-threatening prognosis. A widely disseminated argument against the use of BCG as a therapy for Covid-19 is that BCG induced immune reaction may aggravate the cytokine storm associated with Covid-19 and worsen Covid19 prognosis [16-17]. We therefore monitored Ferritin, and evaluated IL-6 levels, TNF-α and IFN-γ levels in patients on day 0, 7 and 15 after enrollment. As shown in Fig 4A&B, median levels of ferritin over 15 days in SoC group was 447.5 (187.1-656.2) ng/ml while in BCG+SoC group over the same period was 381.6 (180-551.5) ng/ml (p = 0.083, Wilcoxon signed rank). Subjects in SoC show an increase in ferritin (>500 ng/ml) from day 1-8 after enrollment, BCG controls this increase and a spike is seen on day 8-9 after enrollment. In corroboration with Liu et al, (2020) [15], high ferritin levels were also observed in ICU admitted patients in our study (Supp table S4). There was no significant increase in serum levels of IL-6 or TNF-α levels in BCG+SoC group over the SoC group (Fig 4C-F). We also estimated IFN-γ, but did not see detectable levels of IFN-γ in these samples (data not shown).

**Figure 4:**
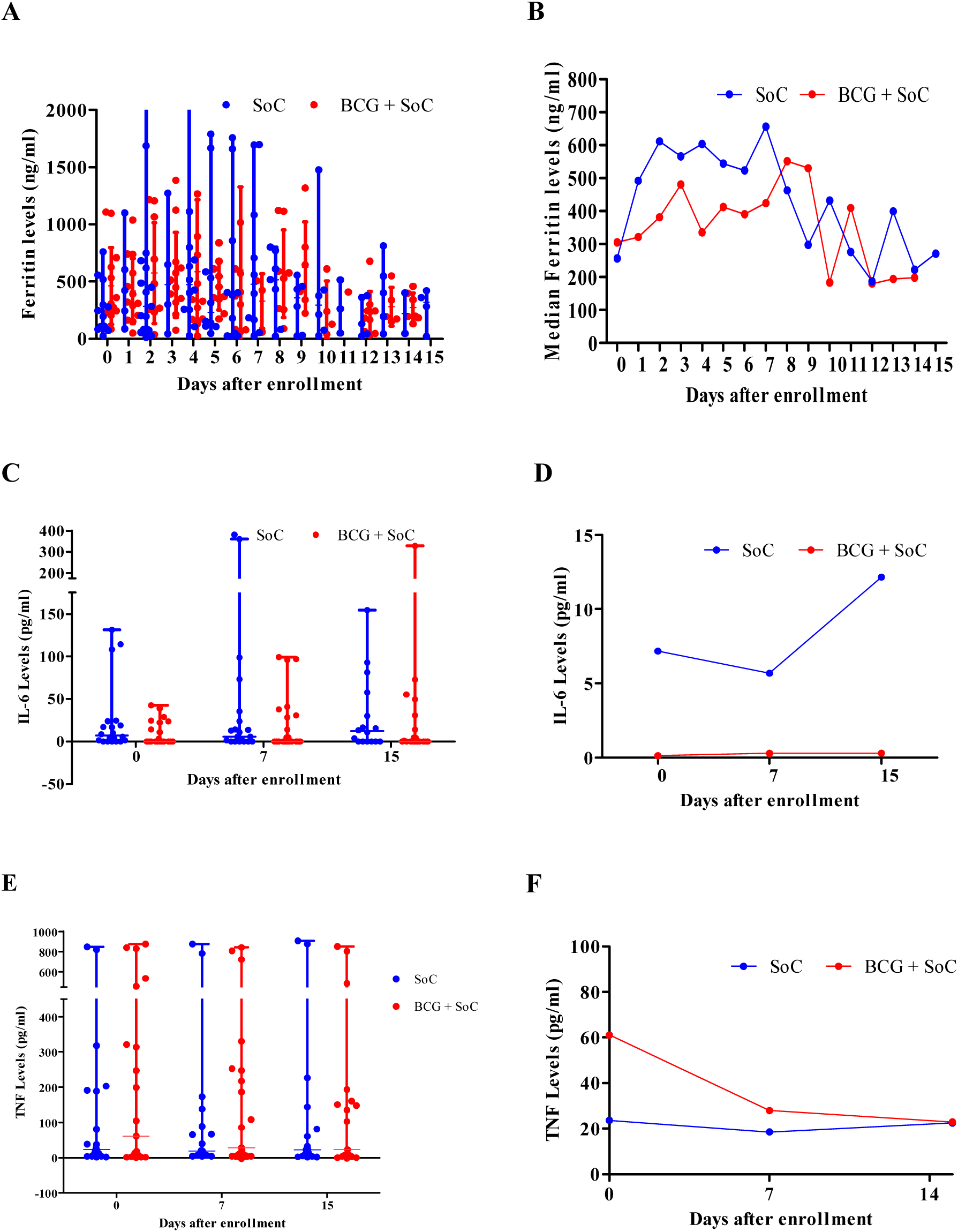
Effect of BCG administration on cytokines. A&B: Ferritin levels (ng/ml). (A) Each point represents ferritin measurements of subjects on the corresponding day. Horizontal bars represent the median values and vertical bars represent min-max ranges of medians calculated for that corresponding day. (B) Each point is the median ferritin levels for 8-16 subjects on that corresponding day. **C&D: IL-6 levels (pg/ml)**. (C) Each point indicates IL-6 levels in serum of each subject on that corresponding as measured by ELISA. Horizontal Bars represent median values, Vertical bars show Min-Max range of values. (D) Each point is the median IL-6 levels for all 30 subjects each subject on that corresponding day. **E&F: TNF-**_α_ **levels (pg/ml)**. (E) Each point indicates TNF-_α_ levels in serum of each subject on that corresponding as measured by ELISA. Horizontal Bars represent median values, Vertical bars show Min-Max range of values. (F) Each point is the median TNF-_α_ levels for all 30 subjects each subject on that corresponding day. For details see Supplementary Appendix.

A comparison of biochemical parameters of all the patients inclusive of serum sodium, potassium, Alkaline phosphatase, SGOT, SGPT, Creatinine, Urea, Uric acid, Bilirubin, Total protein and Albumin (Fig S9) revealed significant differences only in 3 parameters sodium, uric acid and bilirubin (Table S3), none of which were outside the normal reference interval.

## Discussion

Our results indicate that subjects receiving BCG+SoC display a significant improvement in oxygen saturation status within 3-4 days and faster resolution of pneumonia of than subjects receiving SoC thus implicating localized effects in the lung and innate immunity. Such nonspecific effects of BCG that provides immune protection through heterologous mechanisms against diseases other than tuberculosis has also been seen immediately after neonatal vaccination [18-20]. Several modes of action have been proposed for these acute non-specific effects of BCG immunization: innate immunity ‘training’ through control on iron homeostasis, induction of receptor expression on NK cells, monocytes so that response to BCG-unrelated antigens is quicker and larger increase in levels of IL-1β, IL-6, IFN-γ and TNF-α are observed which is also accompanied by a shift of metabolism from oxidative phosphorylation to glycolysis [21-23]. Mice studies indicate that infection with BCG results in acute induction of localized lung immunity through expansion of CD4+, CD8+, and other specific T cells subsets by week 4 that produce IL-6, TNF-α, IFN-γ and systemic increase in IFN-γ levels [24,25]. These observations are reason for being apprehensive of a cytokine storm induced by BCG and complicating Covid-19 prognosis.

As our data shows, the BCG+SoC cohort had lower ferritin levels compared to the SoC group indicating a control on inflammation (see Fig 4). Thus it appears that, BCG induced an iron-withholding response, as has been speculated as mechanism of innate immunity mediated non-specific protection in neonates [21,26]. However, systemic IL-6, TNF-α levels did not significantly differ in both arms indicating absence of any cytokine storm. Levels of IFN-γ were below detection limit over the duration of the study (Fig 4) possibly because of the pulmonary zones involved at enrollment, negative correlation of lower IFN-γ levels with lung fibrosis associated with Covid −19 and the fact that interferon response was seen only 4 weeks after BCG administration in a cohort from South India [27-28].

About 20% of the patients in the study were BCG naïve and they were significantly older than vaccinated patients (Fig S2). Notably, there were no BCG related adversities seen in naïve subjects, instead the fastest hyopoxia resolution was in naïve subjects on BCG+SoC arm whereas in naïve patients on the SoC arm, hypoxia did not improve even after 12-14 days. Comparison between hypoxia resolution in vaccinated and naïve patients on the SoC arm indicates that BCG vaccination in childhood may have some residual effect in offering protection to pneumonia (see Fig 1A-D). Of note, BCG-naïve patients (median age 54.5 yrs) on the SoC arm mimic elderly populations in countries with no or limited BCG vaccination policy where Covid-19 associated severity of disease and mortalities were high.

Indirect assessment of viremia through evaluation of C_T_ values before and after 15th Jun 2020, show that BCG could decrease viral load by day 7. Such a reduction in viral titers on BCG administration have also been seen in case of other viruses like respiratory syncytial virus, influenza A virus and herpes simplex virus type 2 possibly through macrophage activation [29-31]. However this sensitivity was not seen in later half of the study indicating that perhaps there were 2 strains of the virus as suggested by Paul et al. [32]. Sequencing the virus strains is underway at present which will make this clear and could possibly provide an explanation of the heterogeneous nature of Covid-19 progression and outcomes in various populations and/or in the same population /center at different times.

## Conclusions

Primary and secondary outcome analyses clearly indicate better resolution of Covid-19 in patients receiving BCG along with concomitant SoC, with reduction of hypoxia and pneumonia, lesser ICU admissions and deaths, better control of inflammation and an increase in nSARS-2 specific IgG antibodies.

Currently as per the BCG world atlas, 80% of neonates across the globe are vaccinated with BCG [33] with the duration of immunity conferred by BCG varying from 10-20 yrs. Our study suggests that re-vaccination with BCG in adults is safe since we did not see any BCG related adversities in the study population. BCG may also provide additional long-term benefits of risk reduction for tuberculosis and lung cancer [34-35].

To our knowledge this is the first study of the use of BCG as a therapy for Covid-19 that provides a direct comparison between BCG naïve and vaccinated subjects. This is an important takeaway of the study and should pave the way for countries without BCG in their national health policies to introduce BCG as a therapy for Covid-19.

## Data Availability

All data referred to in the manuscript will be provided after due permission from the State Government of Maharashtra after publication and submission to the Drug Controller General of India. Data when provided will be on a case to case basis and for research purposes only.

## Acknowledgements

The authors are grateful to Dr. Sonali Salvi who is the PI of Trial as designated by DGCI but not an author as per her written request, Dr. Dasani, HOD, Pharmacology, & Dr. Harish Tathia, Dept. of Forensic medicine BJMC &SGH for their co-operation and the residents Dr. Subham More, Dr. Shreeja Roy and Dr. Deepak Nair, BJMC &SGH, Pune for their co-operation in monitoring the subjects of this study. The authors are also grateful to Dr. Shashikant Vaidya for his co-operation, Mr. Samir Patil, Mr. Pravin Shirsat who carried out the in vitro analyses for this study and Mr. Bhagyawan Pawar who helped out extensively at HITRT, Mumbai.

## Author Contributions

UP was responsible for the concept, protocol, securing all permissions for the trial in vitro analyses, statistical analyses and paper writing. SS was the on-site physician and responsible for on-clinical site implementation of the trial. Both UP and SS were responsible for study design, analyses and interpretation of the data. RB and SJ were clinical site co-ordinators. SM and RD were responsible for overall co-ordination and permissions for the trial. All authors were responsible for drafting and editing the intellectual content of the manuscript.

