## Supplemental info for "Phase II Clinical trial for Evaluation of BCG as potential therapy for COVID-19"

#### Indian Study

**Table of Contents**


| Sr. No | Description | Page. no |
|---|---|---|
| 1. | Figure S1: Consort Flow Diagram of the Study | 3 |
| 2. | Figure S2: Blinding and randomization | 4 |
| 3. | Standard of Care & Figure S3: Standard of Care analyses | 5 |
| 4. | Figure S4: Analyses of age of subjects in the study | 6 |
| 5. | Table S1: Statistical analyses of SpO2/FiO2 of Figure 1B | 7 |
| 6. | Table S2: Statistical analyses of SpO2/FiO2 of Figure 1C | 8 |
| 7. | Figure S5: Localized reaction to BCG | 9 |
| 8. | Supplementary Methods | 10 |
| 9. | Figure S6: RT-PCR analyses to estimate viremia | 11 |
| 10. | Figure S7: Standard Curves for estimation of IL-6 , TNF-alpha, IFN-gamma and IgG by ELISA | 12 |
| 11. | Figure S8: Analyses of more CBC parameters | 13 - 14 |
| 12. | Figure S9: Analyses of Biochemical parameters | 15 - 18 |
| 13. | Table S3: Effect of BCG on Biochemical parameters | 19 |
| 14. | Table S4: Comparison of Adverse Events in the study | 20 |
| 15. | Final approved protocol from Drug Controller General of India (DGCI) | 21 - 44 |

**Figure S1: CONSORT 2010 Flow Diagram of**

**Phase II Clinical Trial For use of BCG as a potential therapy for Covid-19**

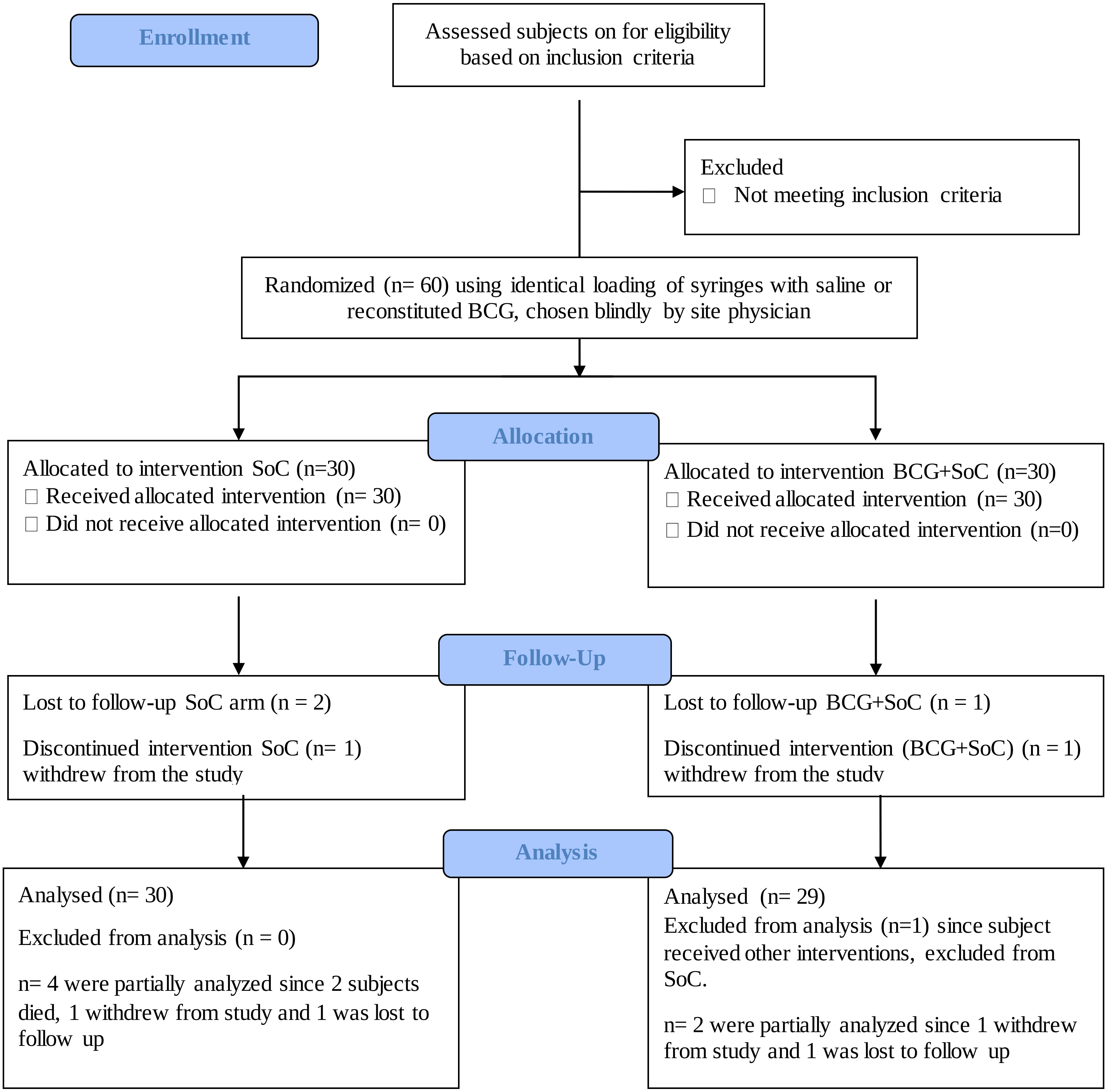

**Figure S2**

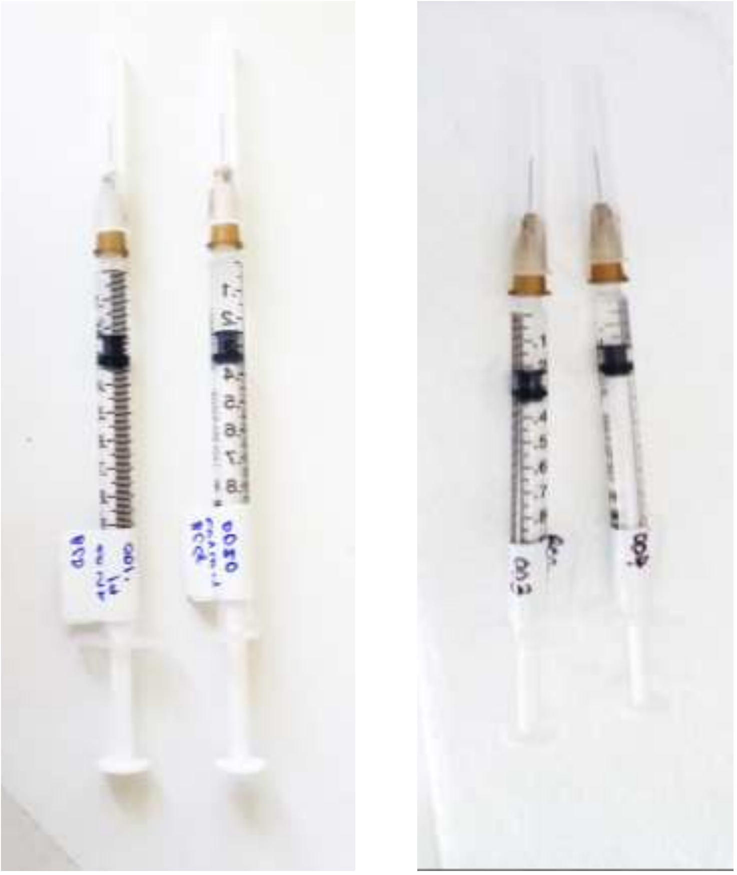

**Figure S2: Blinding and randomization**.

Figure shows identically loaded syringes filled wither with saline or BCG reconstituted in saline. Selection of either one syringe for a screened and enrolled patient ensured that on-site physician and subjects were blinded and randomization was achieved.

**Standard of Care:**

This included the prescribed use of following antibiotics

1. Amoxycillin-clavulanic acid,
2. Piperacillin - tazobactam,
3. Amoxycillin-clavulanic acid followed by Piperacillin - tazobactam,
4. Piperacillin - tazobactam followed by Metronidazole

Isolated cases also received Linezolid or Ceftriaxone in combination with the above.

Apart from the above, all subjects received

1. Hydroxychloroquine
2. Methyl predisolone or dexamethasone
3. Low molecular weight heparin or standard heparin
4. Injection of Pan 40
5. Tablet Vitamin C, Zinc

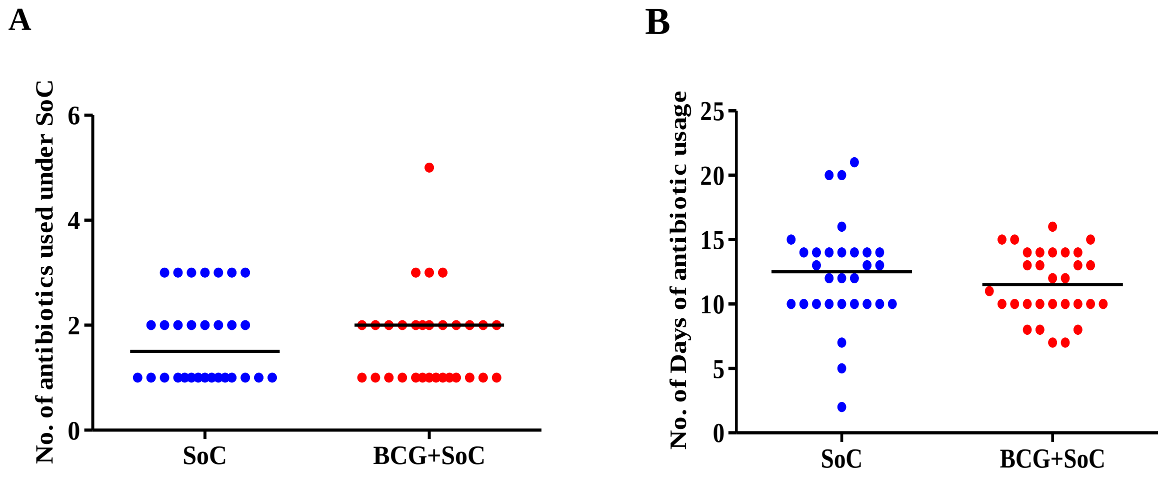


**Figure S3: Comparison of (A) Number of Antibiotics used and (B) Number of days for which it was used between subjects in the SoC and BCG+SoC group.** Each dot corresponds to one patient. Horizontal lines represent median value. Data were compared using T test and Mann whitney analyses.

**Figure S4: Analyses of age of subjects in study**

**A**

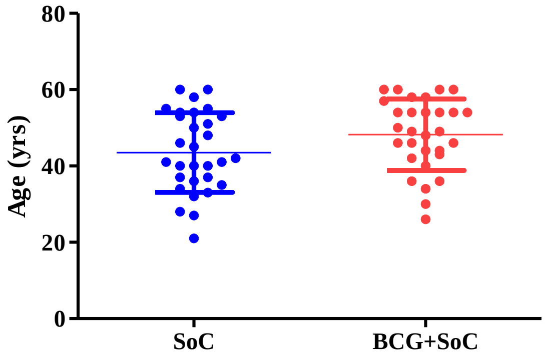


**A**

**Comparison of age of subjects in SoC v/S BCG+SoC arm.** Each dot represents age of one subject. Comparison was carried out using a t test followed by Mann whitney analyses (p = 0.076).

**B**

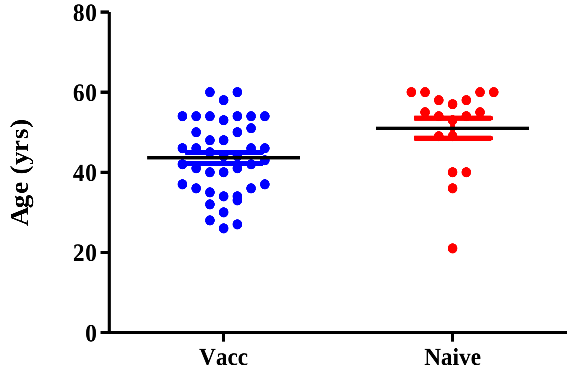


**B. Comparison of age of vaccinated vis a vis naïve subjects.** Each dot represents age of one subject, vaccinated subjects (n=42) and naïve subjects (n = 19). Mean age for Vaccinated 43.64 +/- 9.17 yrs and for Naive 51.06 +/- 10.54 yrs Comparison was carried out using a t test followed by Mann whitney analyses (p = 0.0033).

**C**

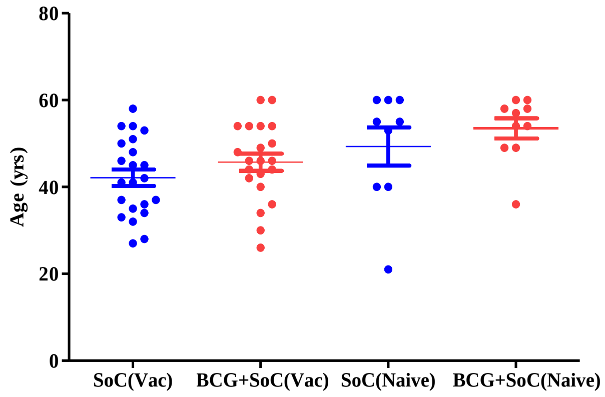


**C. Comparison of age of vaccinated vis a vis naïve subjects in both arms.** Each dot represents age of one subject. Mean age for Vaccinated subjects on SoC (n=22) 42.14 +/- 8.95 yrs, BCG+Soc (n=21) was 45.71 +/- 9.06 yrs, for Naïve subjects on SoC (n=9) was 48.33 +/- 13.21 yrs and on BCG+SoC was 53.50 +/- 7.33 yrs. Comparison was carried out using a t test followed by Mann whitney analyses (p = 0.017).

**Table S1:**

**Linear regression analyses of SpO2/FiO2 data shown in Figure 1 (in main document).**

| **Linear regression analyses** | | |
|---|---|---|
| **Best-fit values** | **SoC** | **BCG+SoC** |
| Slope (per day) | 6.547 ± 1.937 | 13.16 ± 1.219 |
| Y-intercept when X=0.0 | 328.8 ± 13.65 | 309.3 ± 9.630 |
| X-intercept when Y=0.0 | -50.22 | -23.50 |
| 1/slope | 0.1527 | 0.07598 |
| **95% Confidence Intervals** | | |
| Slope | 2.751 to 10.34 | 10.77 to 15.55 |
| Y-intercept when X=0.0 | 302.0 to 355.5 | 290.5 to 328.2 |
| X-intercept when Y=0.0 | -129.0 to -28.36 | -31.24 to -17.83 |
| **Is slope significantly non-zero?** | | |
| F | 11.43 | 116.6 |
| DFn, DFd | 1.000, 261.0 | 1.000, 349.0 |
| **P value** | **0.0008** | < **0.0001** |
| Deviation from zero? | Significant | Significant |
| **Equation** | **y = 6.547(x) + 328.8** | **y = 13.16(x) + 309.3** |

**Table S2:**

**Linear regression analyses of SpO2/FiO2 data shown in Figure 1D (main document).**

| | **SoC (naive)** | **BCG+SoC (naive)** | **Soc (vacc)** | **BCG+SoC (vacc)** |
|---|---|---|---|---|
| **Best-fit values** | | | | |
| Slope | -5.516 ± 1.694 | 13.87 ± 1.048 | 10.68 ± 0.852 | 10.76 ± 1.070 |
| Y-intercept when X=0.0 | 374.6 ± 14.91 | 295.4 ± 9.227 | 313.8 ± 7.502 | 348.1 ± 9.422 |
| X-intercept when Y=0.0 | 67.92 | -21.30 | -29.38 | -32.35 |
| 1/slope | -0.1813 | 0.07212 | 0.09362 | 0.09292 |
| **95% Confidence Intervals** | | | | |
| Slope | -9.149 to -1.882 | 11.62 to 16.11 | 8.853 to 12.51 | 8.466 to 13.06 |
| Y-intercept when X=0.0 | 342.7 to 406.6 | 275.6 to 315.2 | 297.7 to 329.9 | 327.9 to 368.3 |
| X-intercept when Y=0.0 | 43.81 to 184.7 | -26.94 to -17.23 | -37.04 to -23.94 | -43.21 to -25.28 |
| **Goodness of Fit** | | | | |
| $r^2$ | 0.4309 | 0.9259 | 0.9182 | 0.8784 |
| Sy.x | 31.24 | 19.33 | 15.71 | 19.74 |
| **Is slope significantly non-zero?** | | | | |
| F | 10.60 | 175.0 | 157.1 | 101.1 |
| DFn, DFd | 1.000, 14.00 | 1.000, 14.00 | 1.000, 14.00 | 1.000, 14.00 |
| P value | 0.0057 | < 0.0001 | < 0.0001 | < 0.0001 |
| Deviation from zero? | Significant | Significant | Significant | Significant |
| **Equation** | **y = -5.15(x) + 342.7** | **y = 13.87(x) + 275.6** | **y = 10.68(x) + 297.7** | **y = 10.76(x) +327.9** |

**Figure S5**

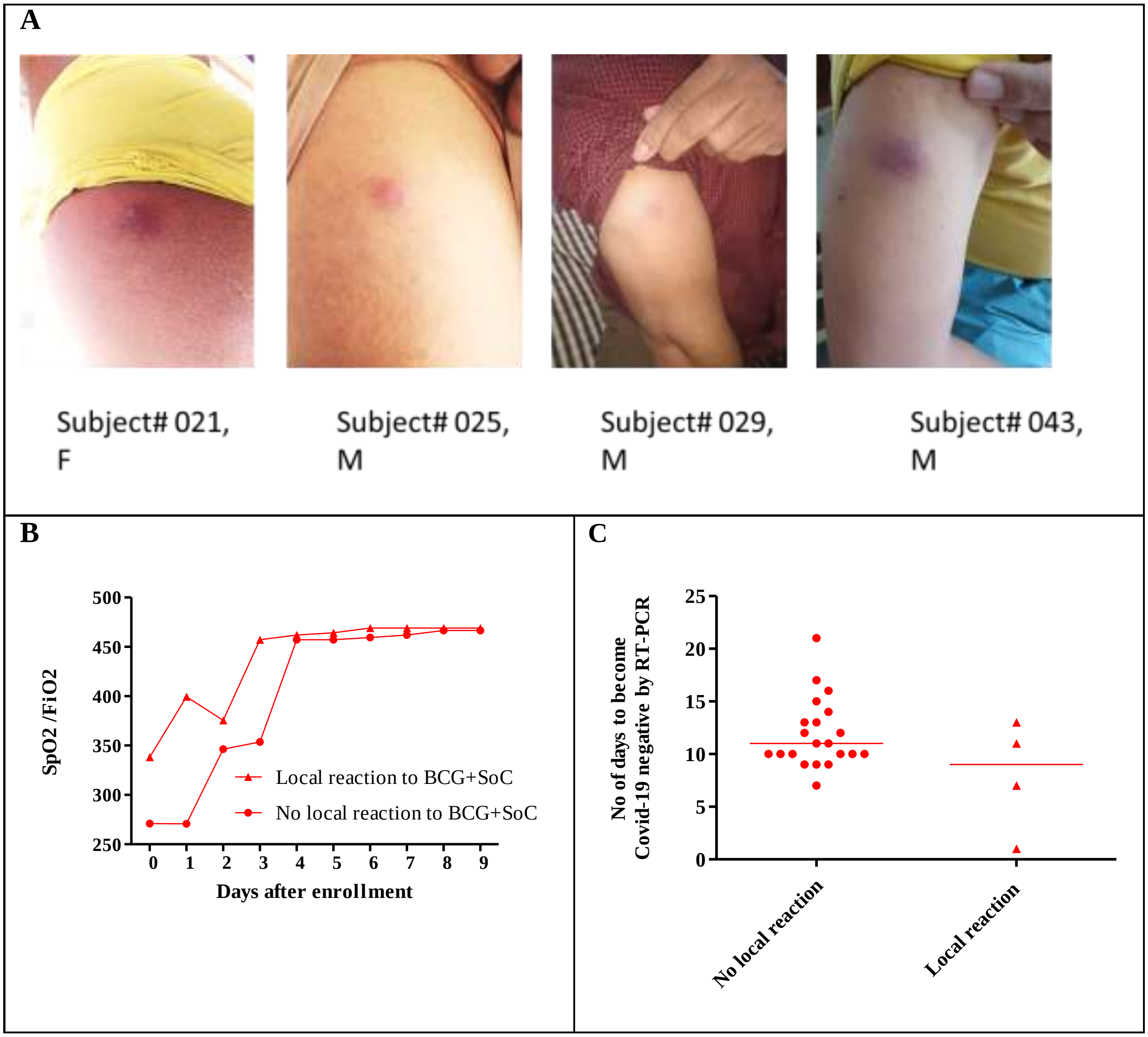


**Figure S5: Localized inflammatory response at the injection site in subjects randomized to the BCG+SoC arm.** A total of n = 4 /60 subjects in the BCG arm showed local inflammatory response after 10 days of receiving BCG. Photographs were taken with permission of subjects.

**Supplementary Methods**

**Evaluation of viremia by Reverse transcriptase- quantitative PCR (RT-qPCR)**

Viral RNA was extracted using RNA extraction kits (Qiagen) from nasopharyngeal swabs of all subjects taken on days 0, 7 and 15 in VTM (HiMedia laboratories / Capricorn) RNA extracted was used to prepare cDNA by incubation with RT, oligo-dT primers and dNTPs at 42 degC for 40 min as per manufacturer's instructions in 10 ul reactions (Takara). Viremia was evaluated using qPCR on a Quant 5.0, Thermo Fisher system. Reactions were 10 uL with 2 uL of cDNA prepared, specific primers for E gene (0.2 uM, each) using a 2x qPCR mix (SyBr green based GoTaq, Promega) using cycling conditions: 95 degC for 2 min followed by 40 cycles of 95 degC 15 sec and 60 degC 1 min. Primers used were

E_Sarbeco_F1 ACAGGTACGTTAATAGTTAATAGCGT

E_Sarbeco_R2 ATATTGCAGCAGTACGCACACA

All reactions were carried out in duplicates. Images of analyses of 60 samples are shown in Supplementary Figure S6

**Estimation of IL-6, TNF-alpha, IFN-gamma, IgM and IgG**

Blood was collected on day 0, day 7 and day 15 of enrollment in heparin vaccutainers (BD Biosciences). Blood was centrifuged at 10,000 rpm for 5 min at RT and plasma was separated. Plasma samples were separated and stored at -80 degC until further analyses.

Estimation of IL-6, TNF-alpha (Krishgen Biosystems), IFN-gamma, IgM and IgG (Raybiotech) levels in plasma was by ELISA. Standard curves were generated using either recombinant human IL-6, TNF-alpha, IFN-gamma standards and positive IgM and IgG samples provided by manufacturers. These standard curves are shown in Supplementary Fig S7.

Undiluted plasma (100 ul) was used in the ELISA assays for IL-6, TNF-alpha, IFN-gamma and IgM levels. For estimation of IgG levels, plasma samples were diluted 1:1000 with sample dilution buffer provided by manufacturer (Raybiotech). All standards and samples were analyzed in duplicates.

**Figure S6**

**A**

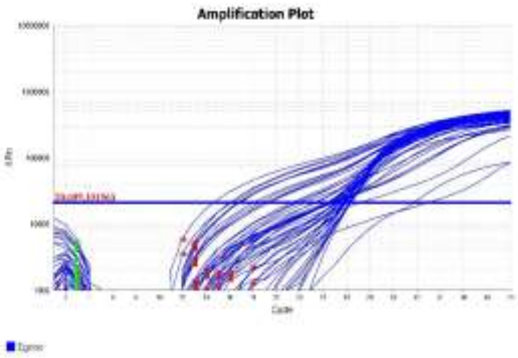


**B**

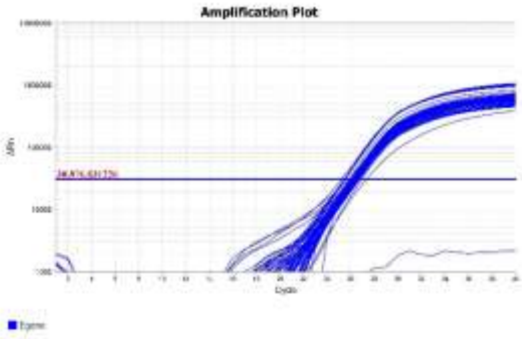


**C**

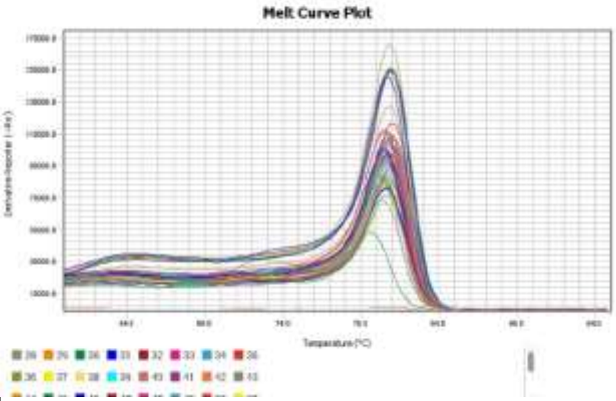


**Figure S6: Estimation of viremia from nasopharyngeal swabs of subs. A & B:** Amplification of cDNA using qPCR: X axes shows the cycle number ($C_T$) and Y axis is the Log of fluorescence (a.u.) signal acquired. Samples in A belonged to the 20 patients who were enrolled in May-Jun 2020 and in B belonged to remaining 40 patients enrolled from Jun-Aug 2020 **C:** Melt curves for E gene of samples corresponding to samples shown above.

**Figure S7**

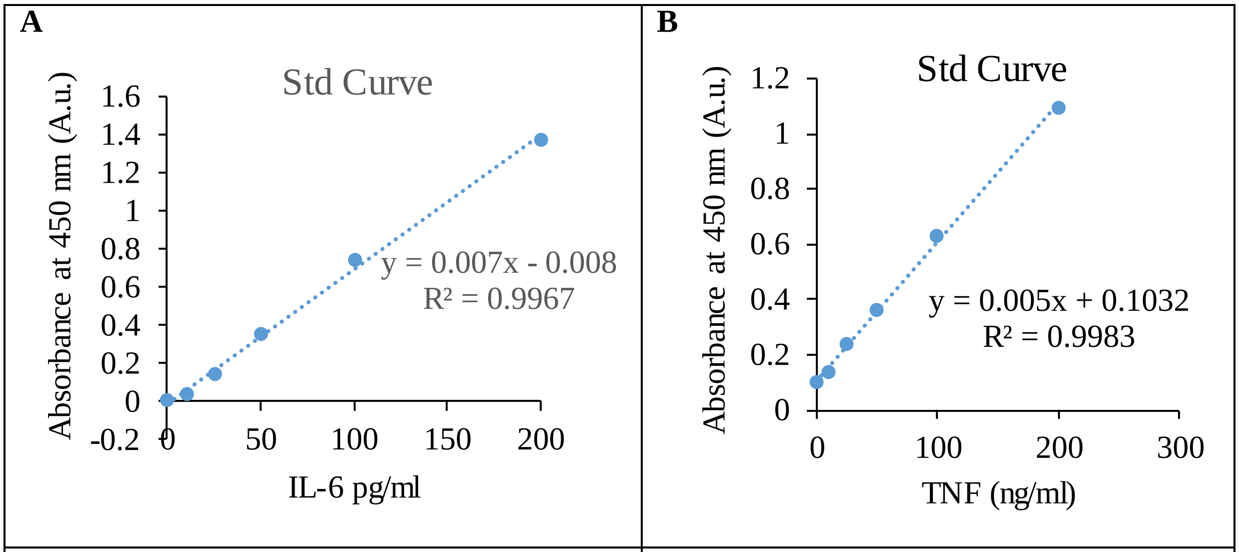


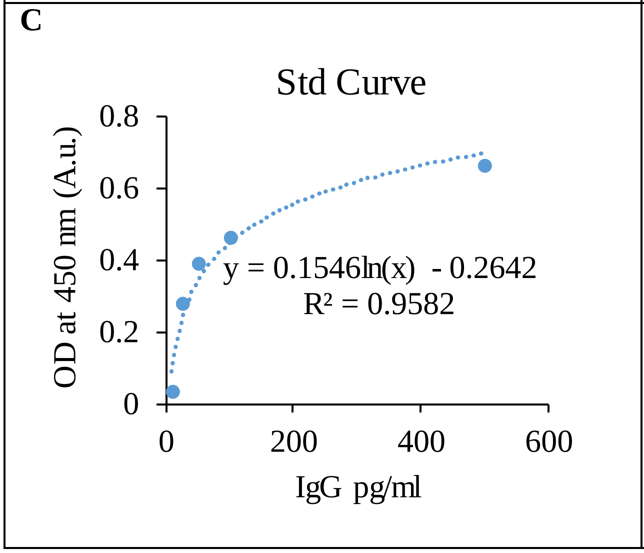


**Figure S7: Standard curves for estimation of (A) IL-6, (B) TNF-alpha and (C) IgG from serum.** Standard curves were generated using either recombinant human IL-6, TNF-alpha, and positive IgG samples provided by manufacturers.

**Figure S8**

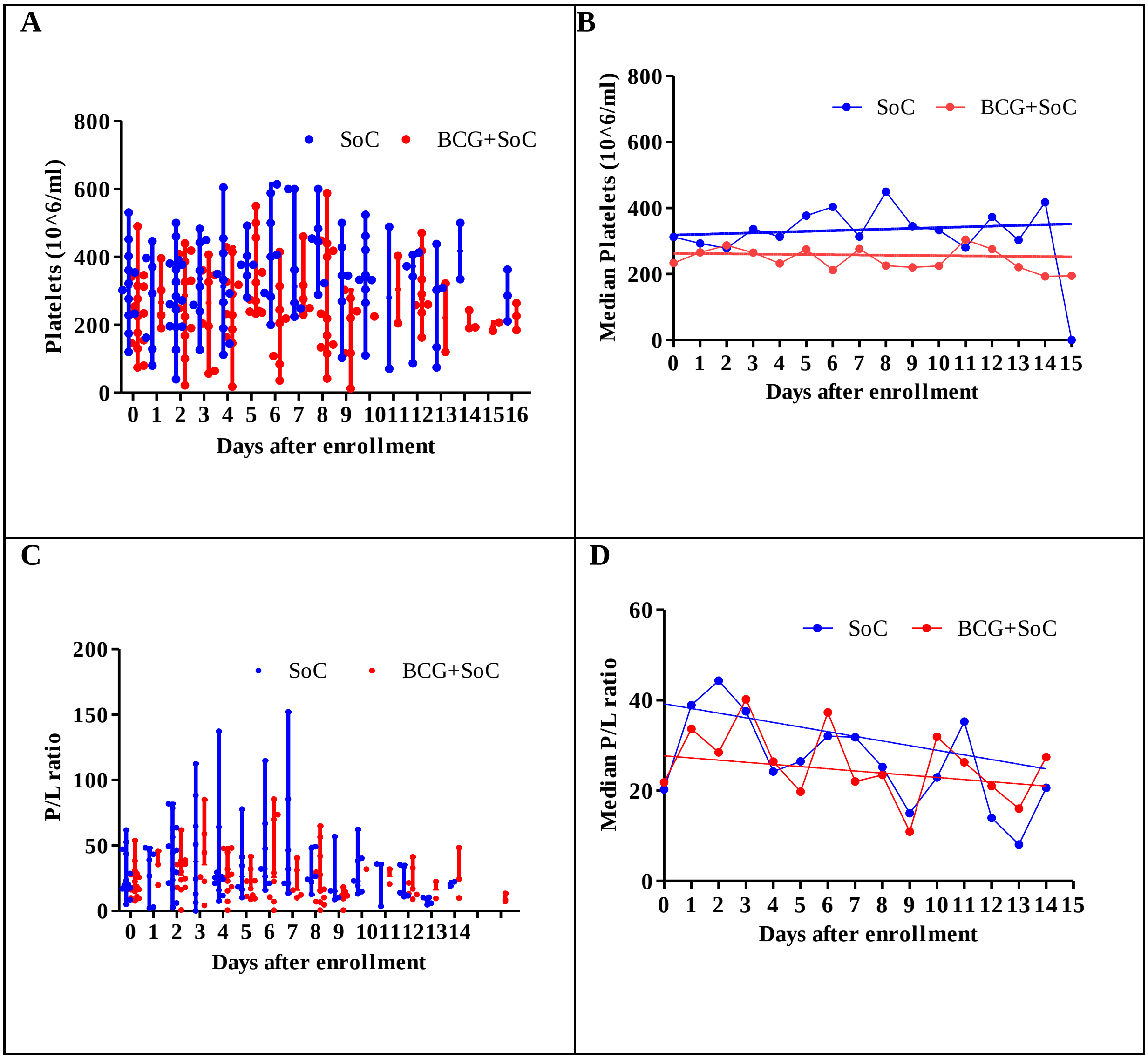
A
Platelets (10^6/ml)
SoC
BCG+SoC
Days after enrollment
B
Median Platelets (10^6/ml)
SoC
BCG+SoC
Days after enrollment
C
P/L ratio
SoC
BCG+SoC
Days after enrollment
D
Median P/L ratio
SoC
BCG+SoC
Days after enrollment

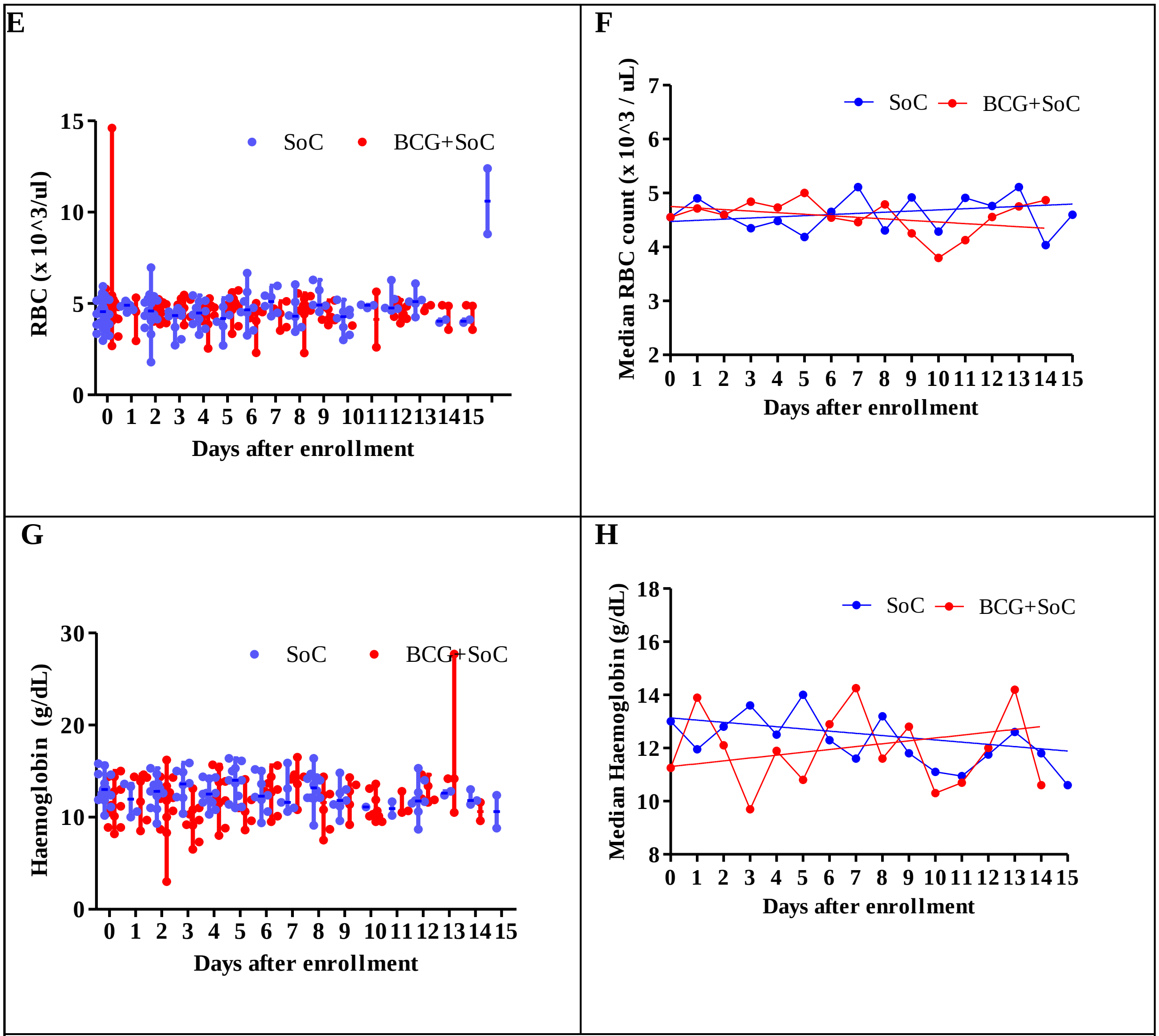


**Figure S8: Effect of BCG on A&B: Platelet count, C&D: P/L ratio, E&F: RBC count, G&H: Haemoglobin level**

**LHS:** Each point represents value of platelets, P/L ratio, RBC count or Haemoglobin level of that subject on the corresponding day. Horizontal bars represent the median values and vertical bars represent min-max ranges of medians calculated for that corresponding day. **RHS:** Each point represents median of platelets, P/L ratio, RBC count or Haemoglobin level of 8-16 subjects on the corresponding day after enrollment. Linear regression analyses was carried out using Graphpad Prism. **P/L ratio:** Platelet to Lymphocyte ratio **SoC**: Standard of Care

**Figure S9**

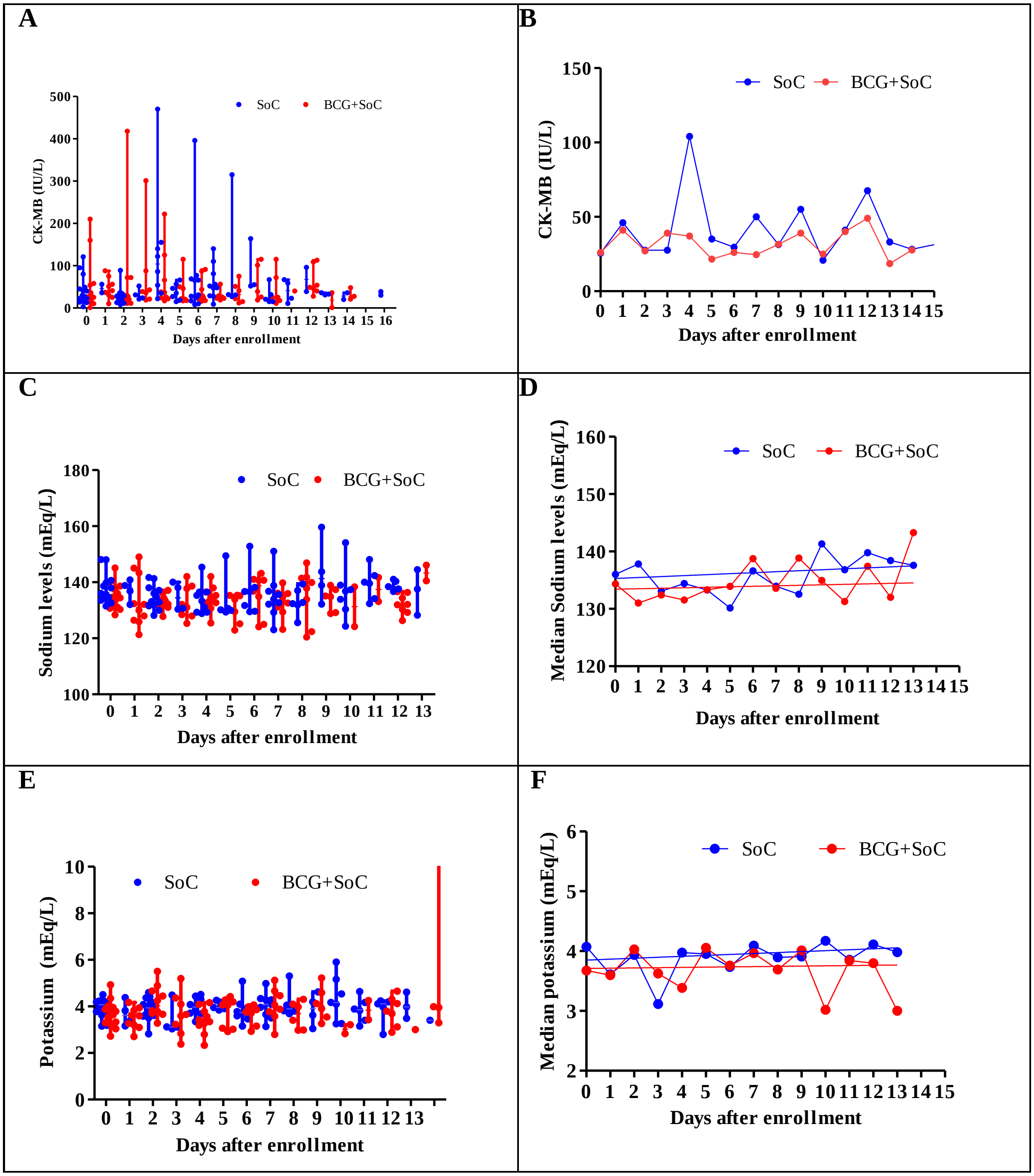
A
CK-MB (IU/L)
SoC
BCG+SoC
Days after enrollment
B
CK-MB (IU/L)
SoC
BCG+SoC
Days after enrollment
C
Sodium levels (mEq/L)
SoC
BCG+SoC
Days after enrollment
D
Median Sodium levels (mEq/L)
SoC
BCG+SoC
Days after enrollment
E
Potassium (mEq/L)
SoC
BCG+SoC
Days after enrollment
F
Median potassium (mEq/L)
SoC
BCG+SoC
Days after enrollment

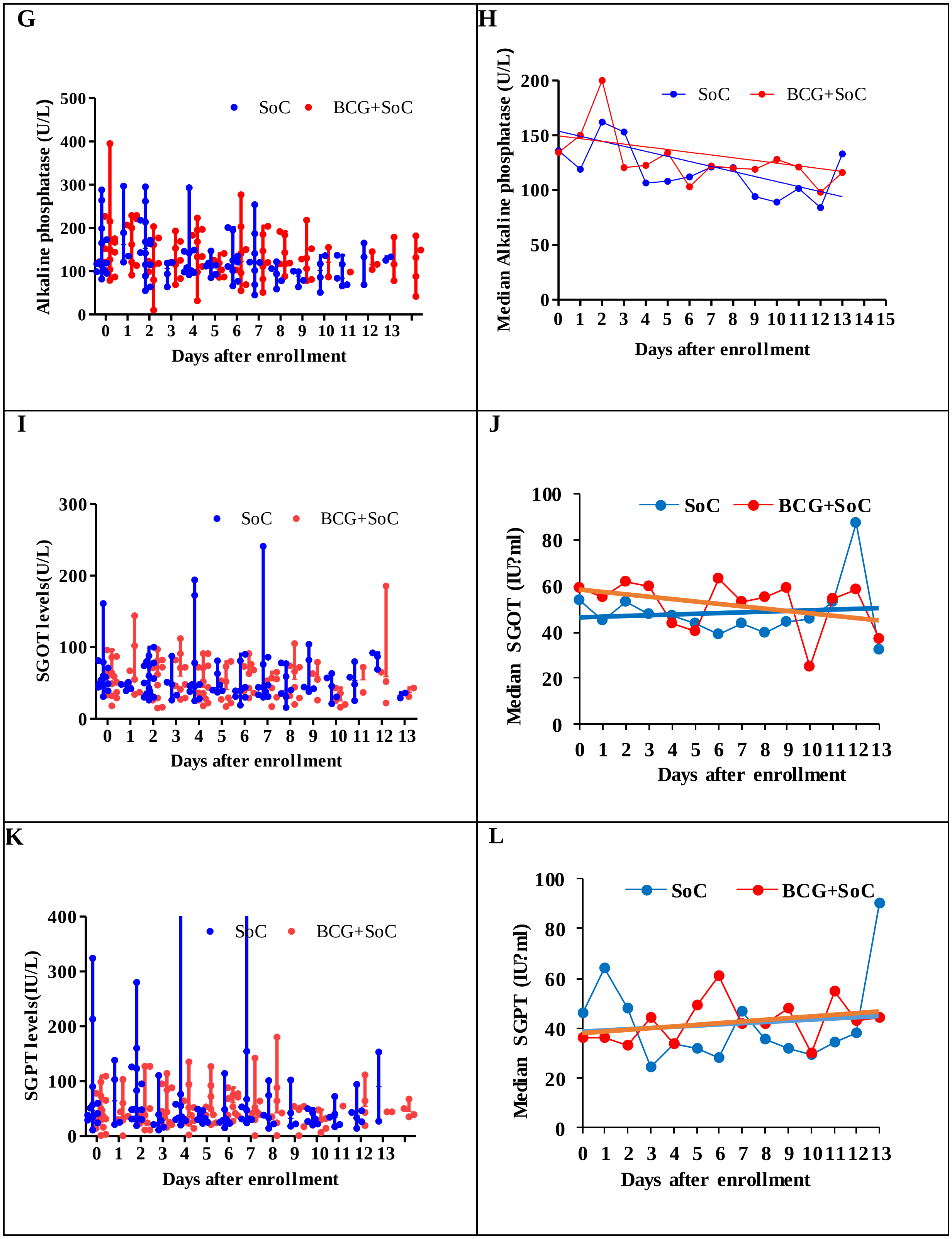
G
Alkaline phosphatase (U/L)
SoC
BCG+SoC
Days after enrollment
H
Median Alkaline phosphatase (U/L)
SoC
BCG+SoC
Days after enrollment
I
SGOT levels(U/L)
SoC
BCG+SoC
Days after enrollment
J
Median SGOT (IU?ml)
SoC
BCG+SoC
Days after enrollment
K
SGPT levels(IU/L)
SoC
BCG+SoC
Days after enrollment
L
Median SGPT (IU?ml)
SoC
BCG+SoC
Days after enrollment

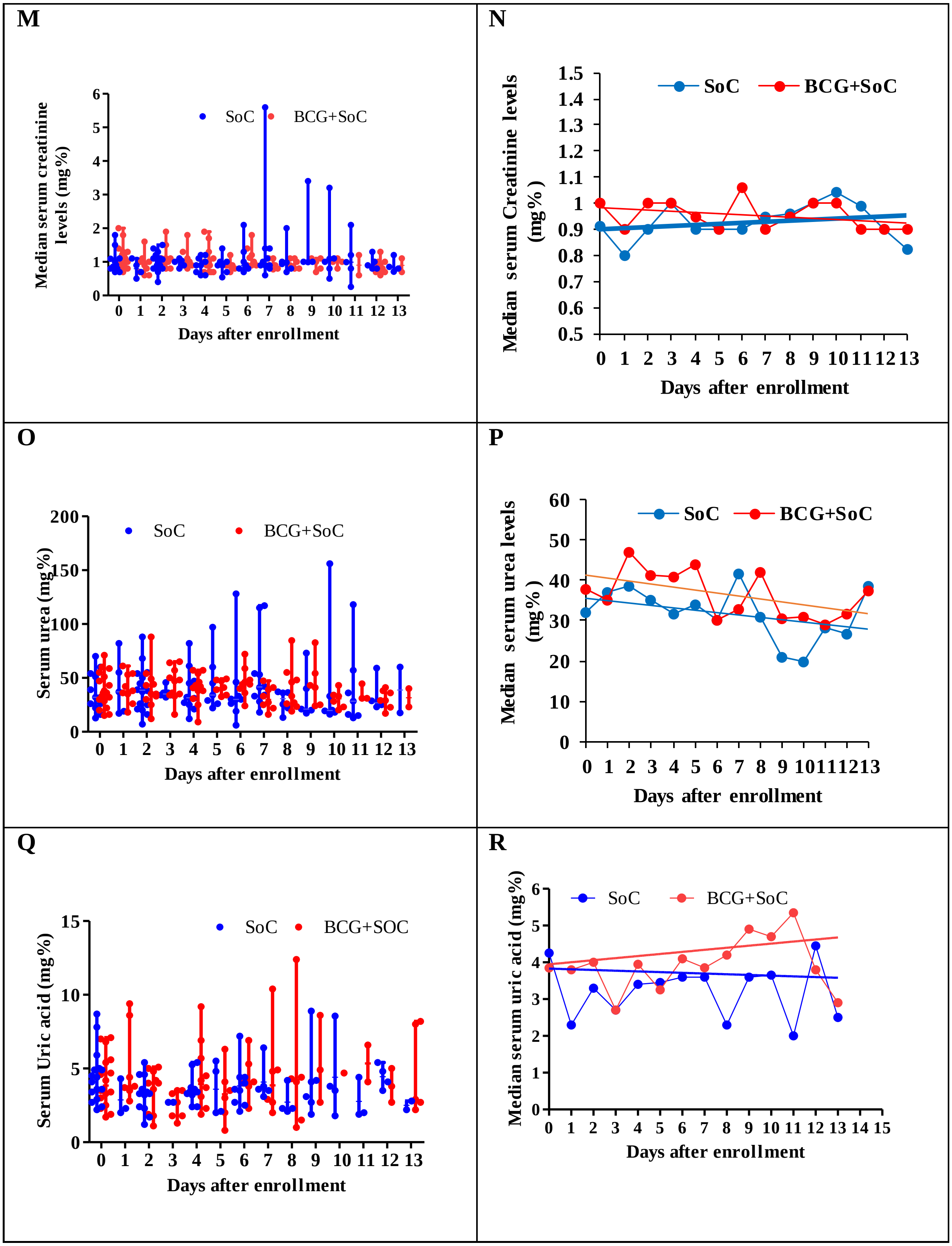
M
Median serum creatinine levels (mg%)
SoC
BCG+SoC
Days after enrollment
N
Median serum Creatinine levels (mg%)
SoC
BCG+SoC
Days after enrollment
O
Serum urea (mg%)
SoC
BCG+SoC
Days after enrollment
P
Median serum urea levels (mg%)
SoC
BCG+SoC
Days after enrollment
Q
Serum Uric acid (mg%)
SoC
BCG+SOC
Days after enrollment
R
Median serum uric acid (mg%)
SoC
BCG+SoC
Days after enrollment

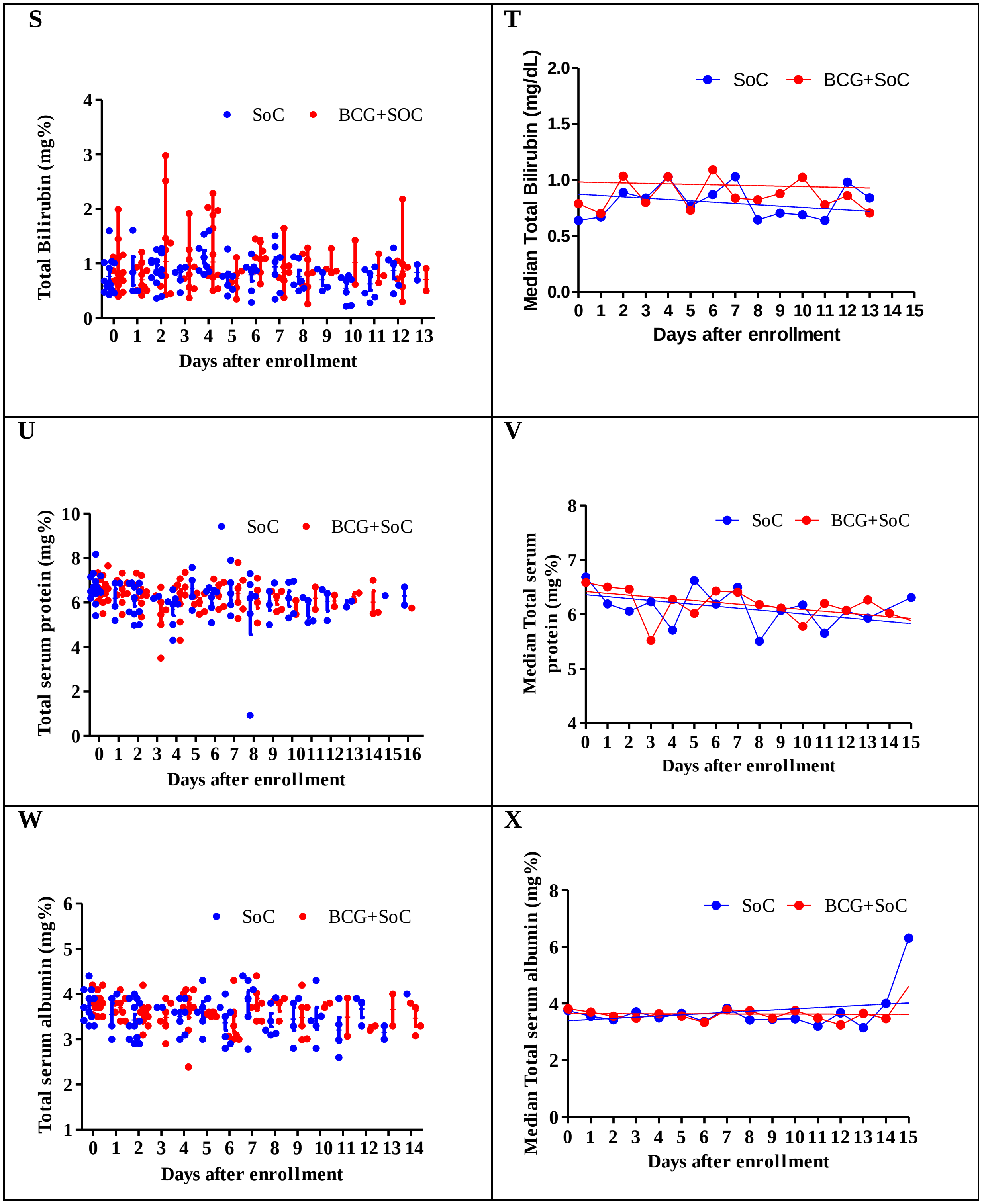
S
Total Bilirubin (mg%)
SoC
BCG+SOC
0 1 2 3 4 5 6 7 8 9 10 11 12 13
Days after enrollment
T
Median Total Bilirubin (mg/dL)
SoC
BCG+SoC
0 1 2 3 4 5 6 7 8 9 10 11 12 13 14 15
Days after enrollment
U
Total serum protein (mg%)
SoC
BCG+SoC
0 1 2 3 4 5 6 7 8 9 10111213141516
Days after enrollment
V
Median Total serum protein (mg%)
SoC
BCG+SoC
0 1 2 3 4 5 6 7 8 9 10 11 12 13 14 15
Days after enrollment
W
Total serum albumin (mg%)
SoC
BCG+SoC
0 1 2 3 4 5 6 7 8 9 10 11 12 13 14
Days after enrollment
X
Median Total serum albumin (mg%)
SoC
BCG+SoC
0 1 2 3 4 5 6 7 8 9 10 11 12 13 14 15
Days after enrollment

**Figure S9: Effect of BCG on serum levels of A&B: CK-MB, C&D: Sodium, E&F Potassium, , G&H: Alkaline phosphtase I&J: SGOT, K&L: SGPT, M&N: Creatinine, , O&P: Urea Q&R: Uric acid, S&T: Bilirubin U&V: Total protein, W&X: Albumin**

**LHS:** Each point represents value of corresponding parameter of that subject on the corresponding day. Horizontal bars represent the median values and vertical bars represent min-max ranges of medians calculated for that corresponding day. **RHS:** Each point represents median of corresponding parameter of 8-16 subjects on the corresponding day after enrollment. Linear regression analyses was carried out using Graphpad Prism. **SoC**: Standard of Care

**Table S3: Effect of BCG on Biochemical parameters (See Supplementary Figure 5)**

| Sr. No | Parameter | p value (Mann Whitney analysis) | Reference interval |
|---|---|---|---|
| 1 | CKMB (U/L) | p = 0.969, not significant | 0 - 24 U/L |
| 2 | Sodium (mEq/L) | p = 0.0180, Median in BCG+SoC lower | 125 - 145 mEq/L |
| 3 | Potassium (mEq/L) | p = 0.1249, not significant | 3.5 - 4.5 mEq/L |
| 4 | Creatinine (mg%) | p = 0.1708, not significant | 0.7 – 1.5 mg% |
| 5 | Uric acid (mg%) | p = 0.0082, Median in BCG+SoC higher | 3.0 - 7.0 mg% |
| 6 | Urea (mg%) | p = 0.981, not significant | 15 - 40 mg% |
| 7 | Bilirubin (mg/dL) | p = 0.0456, Median in BCG+SoC higher | 0.2 - 1.0 mg/dL |
| 8 | SGOT (U/L) | p = 0.963, not significant | 8 - 45 U/L |
| 9 | SGPT (IU/L) | p = 0.777, not significant | 1 - 40 IU/L |
| 10 | Alkaline phosphatase (U/L) | p = 0.418, not significant | 15 - 40 U/L |
| 11 | Total serum protein (gm%) | p = 0.777, not significant | 6 - 8 gm% |
| 12 | Total serum albumin (mg%) | p = 0.745, not significant | 3.5 - 4.5 mg% |

**Table S4: Comparison of Adverse Events in the study**

| **Parameter** | **Subject no. 10** | **Subject No. 50** | **Subject No. 36** | **Subject No. 46** | **Subject No. 17** |
|---|---|---|---|---|---|
| Nature of SAE | ICU admission and death | Death | ICU admission and remission | ICU admission | ICU admission |
| Arm | SoC | SoC | SoC | SoC | BCG+SoC |
| Age | 54 | 51 | 53 | 36 | 60 |
| Gender | Male | Male | Male | Female | Female |
| BCG vaccination status | Vaccinated | Vaccinated | Vaccinated | Vaccinated | Not vaccinated |
| Co-morbidity | None | None | None | None | None |
| Days of SAE after enrollment | NIV: 7 days Death: 11 days after enrollment | Death: 4 days after enrollment | NIV & ICU: 1 day Out of ICU: 4 days Remdesivir: administered after 6 days | NIV & ICU: 2 days and after 2 days withdrew from study | 4 days to NIV and after 7 days withdrew from study |
| N/L ratio | 26.56 | 9.84 | 22.19 | 2.22 | 8.33 |
| Ferritin at admission | 1214 ng/ml | 604.9 ng/ml | 1101 ng/ml | 596 ng/ml | 1039 ng/ml |
| IL-6 at admission | 17.0 pg/ml | n.d (sample hemolysed) | 131.35 pg/ml | n.d (sample hemolysed) | 22.0 pg/ml |

### FINAL APPROVED PROTOCOL FOR CONDUCTING CLINICAL TRIAL

#### PHASE 2 CLINICAL TRIAL FOR THE EVALUATION OF BCG AS POTENTIAL THERAPY FOR COVID-19

2] Table of Contents


| Sr. No | Documents to be submitted | Status | Page. No |
|---|---|---|---|
| | | Yes / No | |
| 1. | Title Page | Yes | 5-6 |
| 2. | Protocol Title, Introduction | Yes | 7-9 |
| 3. | Study Objective(s) (Primary as well as secondary) and their logical relation to the study design. | Yes | 10 |
| 4. | Study Design | Yes | 11-12 |
| 5. | Study Population | Yes | 13 |
| 6. | Study Eligibility - Inclusion & Exclusion Criteria | Yes | 13 |
| 7. | Study Assessments | Yes | 14 |
| 8. | Study Treatment | Yes | 16-17 |
| 9. | Adverse Events | Yes | 18 |
| 10. | Ethical Considerations | | |
| 11. | Study Monitoring & Supervision | Yes, Submitted | 20 |
| 12. | Investigational Product | Yes | 21 |
| 13. | Data Analysis | Yes | 22 |
| 14. | Undertaking by the Investigator | Yes, Submitted | 23 |
| 15. | Informed Consent Documents | Yes, Submitted | 23 |
| 16. | Undertaking by the Sponsor/Sponsors | Not applicable, since it is Govt. Trial<br>Insurance Appendix 5 | 23 |
| 17. | Declaration regarding financial status of the applicant | Not applicable, since it is Govt. Trial | 23 |
| 18. | List of Investigators including site address (es). | Yes | 23-24 |
| 19. | Ethics Committee approvals, if available:- (Institutional Ethics Committee should be in same area where the site is located) | Yes registration of Ethics attached<br>Ethics approval attached | 24 |
| 20. | As per the protocol, whether the subjects will receive the standard care. (Give declaration) | Not applicable, since it is Govt. Trial | 25 |

| | | | |
|---|---|---|---|
| 21. | Details of the contract entered by the sponsor with the investigator/institutions with regard to financial support, amount of fees, honorarium, payments in kind etc. to be paid to the investigator.<br>In case no contract has yet been entered with any Investigator / Institution, plan for financial support, fees, honorarium, and payments in kind etc. to be paid to the investigator | MoU signed with<br>BJ Medical College<br>Attached | 25 |

**(a) Full title of the clinical study**

***Phase 2 Clinical Trial for the Evaluation of BCG as potential therapy for COVID-19***

**(b) Protocol/Study number and protocol version number with date**

*Version 1.0, dated 17.04.2020*

**(c) The IND name/number of the investigational drug**

BCG / Brand Name: Tubervac (Serum Institute of India)

**(d) Complete name and address of the Sponsor and contract research organization if any**

Medical Education & Drugs Department, Government of Maharashtra, 9th floor Mantralay, GT Hospital Campus, Fort, Mumbai 400 001.

Haffkine Institute for Training, Research & Testing, Acharya Donde Marg, Parel, Mumbai 400 012

**(e) List of the Investigators who are conducting the study, their respective institutional affiliations and site locations**

***Overall co-ordinator***

Dr. Sanjay Mukherjee, Hon. Prin Secretary, Medical Education & Drugs Department, 9th floor Mantralay, GT Hospital Campus, Fort, Mumbai 400 001.

Dr. Rajesh Deshmukh, Director, Haffkine Institute for Training, Research & testing, Acharya Donde Marg, Parel, Mumbai 400 012

***Scientific***

Dr. Usha Padmanabhan, Haffkine Institute for Training, Research & Testing, Acharya Donde Marg, Parel, Mumbai 400 012

Dr. Shashikant Vaidya, Haffkine Institute for Training, Research & Testing, Acharya Donde Marg, Parel, Mumbai 400 012

***Scientific***

Dr.Sonali Salvi, BJ Govt. Medical College & Sassoon General Hospital, Jayprakash Narayan Road, Near Pune Railway Station, Pune 411001

Dr. Rohidas Borse, BJ Govt. Medical College & Sassoon General Hospital, Jayprakash Narayan Road, Near Pune Railway Station, Pune 411001

Dr. Samir Joshi, BJ Govt. Medical College & Sassoon General Hospital, Jayprakash Narayan Road, Near Pune Railway Station, Pune 411001

Dr. Sanjay Gaikwad, BJ Govt. Medical College & Sassoon General Hospital, Jayprakash Narayan Road, Near Pune Railway Station, Pune 411001

Dr. Harish Tatia, BJ Govt. Medical College & Sassoon General Hospital, Jayprakash Narayan Road, Near Pune Railway Station, Pune 411001

Dr. Yogesh Gawali, BJ Govt. Medical College & Sassoon General Hospital, Jayprakash Narayan Road, Near Pune Railway Station, Pune 411001

**(f) Name(s) of clinical laboratories and other departments and/or facilities participating in the study.**

BJ Govt. Medical College & Sassoon General Hospital, Jayprakash Narayan Road, Near Pune Railway Station, Pune 411001

Haffkine Institute for Training, Research & Testing, Acharya Donde Marg, Parel, Mumbai 400 012

#### Evaluation of BCG as potential therapy for COVID-19

**Background and Introduction.**

The novel coronavirus nCoV-19 (or SARS-CoV-2 or 2019-nCoV), responsible for the global pandemic COVID-19 was isolated from human airway epithelial cells from patients from Wuhan, China in December 2019 (Wang et al, 2020; Zhu et al, 2020). Seven coronaviruses (CoVs) have been described so far infecting humans of which the SARS-CoV (Kuiken et al, 2003), MERS-CoV and nCoV-19 are serious threats to humans. No vaccines or therapies have been approved for SARS or MERS thus far, demonstrating the need to develop effective therapy or vaccines.

*BacilleCalmette-Guérin*, BCGis a vaccine against tuberculosis that is prepared from a strain of the attenuated (weakened) live bovine tuberculosis bacillus, *Mycobacterium bovis*. The bacilli have retained enough strong antigenicity to become an 80% effective vaccine for the prevention of human tuberculosis.

Overall, BCG vaccine reduces the risk of pulmonary and extra-pulmonary tuberculosis (TB) by approximately 50%, but it has 64% efficacy against TB meningitis and 78% against disseminated TB disease. BCG vaccine also provides some protection against leprosy and non-tuberculous mycobacterial infections. In addition, it has been used in the treatment of superficial carcinoma of the bladder.

.

India and Pakistan introduced BCG mass immunization in 1948, the first countries outside Europe to do so. BCG as a vaccine is safe to be used in children within a week of their birth and is in the Universal immunization programs of many countries in South East Asia and Africa. It has been shown to reduce severe respiratory distress in children from Africa and conferred beneficial immunity and favorable outcomes to malarial infections.

Revaccination with BCG has been tried in some populations (Japanese adults). However the longevity of immune protection due to re-vaccination has not yet been confirmed.

Currently, five main strains account for more than 90% of the vaccines in use worldwide with each strain possessing different characteristics. The strains include the Pasteur 1173 P2, the Danish 1331, the Glaxo 1077 (derived from the Danish strain), the Tokyo 172-1, the Russian BCG-I, and the Moreau RDJ strains (Hayashi et al, 2009).

BCG is known to induce a potent Th1-type response to Mycobacterium antigens and promote the production of both Th1- and Th2-type cytokines in response to unrelated vaccines. Most studies in school children of Brazil and Japan immunized with BCG show a 10-15 yr immunity period conferred by BCG

Management of local BCG complications (injection site reactions and suppurative or non-suppurative lymphadenitis) varies between clinicians, and the optimal approach remains uncertain. Widespread use of the BCG has demonstrated some advantages, such as excellent immune adjuvant activity, long-persisting effects, safety, and low cost.

**542 Drugs and Cosmetics Rules, 1945**

***Relevant information regarding pharmacological, toxicological and other biological properties of the drug/biologic/medical device and previous efficacy and safety experience should be described.***

Each strain of BCG has a different reactogenicity profile - The Pasteur 1173 P2 and Danish 1331 strains are known to induce more adverse reactions than the Glaxo 1077, Tokyo 172-1, or Moreau RDJ strains (Hayashi et al., 2009). The strain is one of the important factors that has been implicated in incidence of adverse events following BCG vaccination (Milstienet al, 1990, Lotte et al.,1984). The BCG to be used in this protocol is Tubervac (Serum Institute of India) is derived from the Russian strain, also known as Moscow strain.

***Previous clinical work with the new drugs should be reviewed here and a description of how the current protocol extends existing data should be provided. If this is an entirely new indication, how this drug was considered for this should be discussed.***

Miller et al, 2020 show a negative correlation between BCG immunization status of a country and mortalities due to COVID-19. In particular, Miller et al., 2020 have presented epidemiological data,that suggests that BCG could be effective against nCoV-19 or SARS-CoV-2. The data (yet to be peer reviewed) found that countries that do not have a BCG immunization policy have more COVID-19 deaths and cases. These countries include the US, the Netherlands and Italy. Countries like Iran which started giving the vaccine late in 1984, had high mortality, suggesting that BCG protected the vaccinated elderly population, whereas countries like Japan have reported lesser cases and mortalities.

Two international trials are on for assessing BCG as a prophylactic agent in healthcare workers in Australia and Netherlands against COVID-19.

It is likely, however, that BCG stimulates general immune response. This results in faster response to infections that could reduce severity of disease and lead to faster recovery. There does not appear to be a direct evidence that BCG will work against nCoV-19 or SARS-CoV-2. It is the purpose of this study to investigate a direct link between BCG intervention and favorable outcome for COVID-19.

**Study Rationale**

WHO and all experts acknowledge that vaccination with BCG leads to general immune boosting with Interferon-gamma being the key player in BCG mediated immune response. BCG is used in the immunization programs largely because of its capacity to resolve tuberculosis, although its role in treating pulmonary tuberculosis remains controversial.

Based on Report of the WHO-China Joint Mission on Coronavirus Disease 2019 (COVID-19) i.e. 55924 laboratory confirmed cases,

Typical signs and symptoms of COVID 19 positive symptoms include:

| | | |
|---|---|---|
| fever (87.9%), | dry cough (67.7%), | fatigue (38.1%), |
| sputum production (33.4%), | shortness of breath (18.6%), | sore throat (13.9%), |
| headache (13.6%), | myalgiaor arthralgia (14.8%), | chills(11.4%), |
| nausea or vomiting (5.0%), | nasal congestion (4.8%), | diarrhea (3.7%), |
| hemoptysis (0.9%), | conjunctival congestion (0.8%) | |

It is interesting to note that Interferon-gamma treatment was also proposed as a possible therapy, for Covid-19, however it was discarded due to potential pulmonary toxicity associated with IFN-gamma.

Wrt to this protocol, hospitalized subjects symptomatic of Covid-19 with fever, dry cough and breathlessness will be potential subjects for the trial. Confirmation of enrollment in study will be after positive RT-PCR report.

Our data analyses shows that in India COVID-19 maximally affects the age group of 20 - 50 yrs. Hence the age group of our subjects are 20 - 50 yrs.

The total number of subjects is kept at 20 (~ 10% of hospitalized cases in local area).

Covid -19 resolves in 2 weeks for mild cases and from 3-6 weeks for severe conditions. It takes an average of 8 weeks for mortality to set in. Hence the total study duration is kept at 3 months (12 weeks).

The kinetic time frame of COVI-19 is comparable or longer than reported for BCG induced immune responses, which is why we believe that BCG may prove to be a cost effective solution to mitigating COVID-19 morbidities.

Therefore we are proposing to use BCG as a therapeutic agent and not as a prophylactic agent.

Given the time period for BCG to induce protective immunity, the trial hopes to see whether the immunity decreases hospitalization of patients with favorable outcomes.

#### 3] Study Objective(s) (Primary as well as secondary) and their logical relation to the study design.

**Main Objective**

The objective of this trial is to evaluate the effects of BCG administration on Covid-19 i.e. disease progression and viremia in patients.

**Primary Outcome Measures:**

1. Total duration of Hospitalization with COVID-19 symptoms such as fever, cough and dyspnea. [Time Frame: from admission until discharge]
2. Decrease in Viral Titer [Time Frame: Measured on day of enrolment, on day 7 and 15 after intervention from nasophayngeal swabs]
3. Duration of COVID-19 symptoms [Time Frame: At time of admission, following enrollment until discharge]

**Secondary Outcome Measures:**

1. No. of ICU admissions [Time Enrolment to 3 months in case of relapse or reinfection]
2. Duration of ICU admission
3. Number of participants needing mechanical ventilation
4. Duration of Mechanical ventilation
5. Mortality [Time From enrolment]
6. Time for resolution of COVID-19 disease [Time From enrolment]
7. Hospitalization cost [Time From enrolment]
8. Change in IFN-gamma, TNF-alpha and IL-6 as evaluated by ELISA [Time Day 0 and days 7 and 15 after BCG intervention]
9. Change in IgG and IgM induced by nCoV-19 in serum [Time Day 0 and days 7 and 15 after BCG intervention]
10. Change in total IgG and IgM levels in serum [Time Day 0 and days 7 and 15 after BCG

intervention]

4] **Study design**

***(a) Overview of the Study Design: Including a description of the type of study (i.e., double-blind, multi-centre, placebo controlled, etc.), a detail of the specific treatment groups and number of study***

**Type of study:**

Randomized double Blind (Subjects in both arms will be given either BCG or saline 0.1 ml from bottles that look identical but will be coded).

**Single centric / Multicentric:** Single centre (patients from 1 center will be enrolled)

**No. of Groups** = 2

**No. of subjects in each group** = 30

**Duration of study:** 3 months.

***(b) Flow chart of the study***

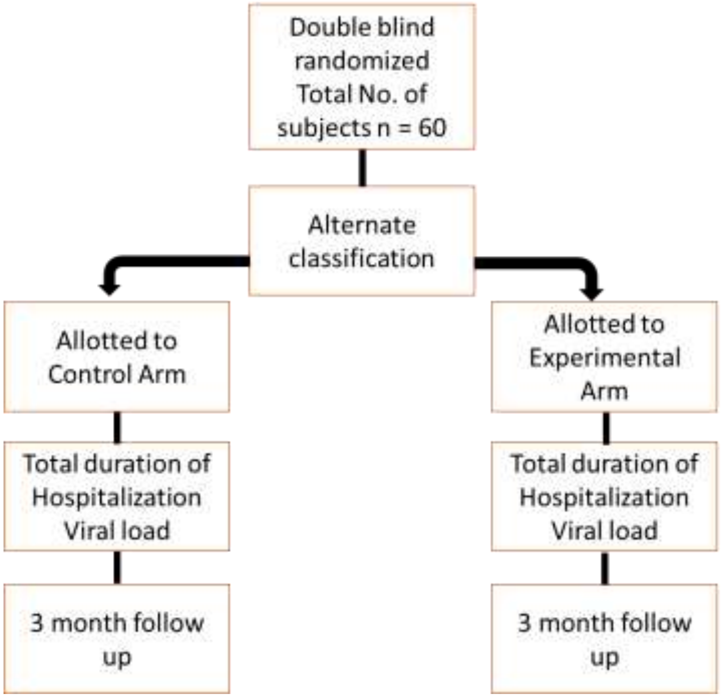

*(c) A brief description of the methods and procedures to be used during the study.*

Subjects with COVID-19 disease defined as

- fever (using self-reported questionnaire), plus
- at least one sign or symptom of respiratory disease including cough, shortness of breath, respiratory distress/failure, runny/blocked nose (using self-reported questionnaire), plus
- positive SARS-Cov-2 test (using RT-PCR as prescribed by WHO, ICMR and NCDC)

*(d) Discussion of Study Design:*

*This discussion details the rationale for the design chosen for this study.*

The time lines chosen for the study are based on the following events

| | |
|---|---|
| Day 0 | Exposure to nCoV-19 |
| Day 0 - 5 | Fever |
| Day 5 - 7 | Typical day of hospitalization |
| Day 7 | IgM production in response to nCoV-19 |
| Da 0 - 14 | Resolution of mild COVID-19 |
| Day 14 - 28 | IgG production in response to nCoV-19 |
| Day 14 - Day 56 | Resolution or deterioration of nCoV associated SARI |
| Day 0 -Day 14 | IFN-gamma production in response to BCG in Japanese adults re-vaccinated with BCG |

5] Study Population

**GROUP I**

**Experimental Group: BCG + Standard of Care treatment**

Participants will receive a single dose of BCG vaccine (Tubervac). The adult dose of BCG vaccine is 0.1 mL injected intradermally over the distal insertion of the deltoid muscle onto the humerus (approximately one third down the upper arm).

**COMPOSITION:** Live, attenuated BCG Vaccine (Bacillus Calmette-Guerin Strain) Each 1 ml contains between 1 x $10^6$ and 33 x $10^6$ Colony Forming Units (C.F.U.)

Diluent: Sodium Chloride Injection I.P.

**ROUTE OF ADMINISTRATION:** Intradermal Injection

**DOSAGE:** BCG should be administered in the dose of 0.1 ml, Intradermal for adult.

**GROUP II**

**Control group: Saline + Standard of Care treatment**

Subjects will receive 0.1 ml saline injected intradermally

#### 6] Subject Eligibility

***(a) Inclusion Criteria.***

Hospitalized subjects with confirmed COVID-19 will be included in this as per following criteria:

1. Age 18-60 years
2. Symptomatic subjects with fever plus at least one sign or symptom of respiratory disease including cough, shortness of breath, respiratory distress/failure, runny/blocked nose , plus
3. Positive SARS-Cov-2 test at admission (using RT-PCR as prescribed by WHO, ICMR and NCDC)
4. Patients with co-morbidities like hypertension will be included after normalization of blood pressure respectively, after assessment by study physician.

***(b) Exclusion Criteria.***

Subjects outside the age group

Subjects who test negative for nCOV-19 by RT-PCR as per criteria laid down by ICMR

Subjects with

1. Any co-morbidities such as acute kidney injury, chronic kidney disease, congestive heart failure, chronic liver disease
2. Active tuberculosis of any organ
3. Diabetes
4. Immunodeficiency disorders – HIV, active Hepatitis B or hepatitis C infection, organ transplant recipient, patients receiving systemic corticosteroid therapy.
5. Systemic lupus erythematosus, rheumatoid arthritis, and other autoimmune diseases
6. Hypogamma-globulinemia,
7. Congenital immunodeficiency disorders
8. Sarcoidosis,
9. Leukaemia,
10. Solid tumor malignant disease with or without metastases
11. Chronic eczema or other generalised dermatological disorder
12. Pregnant women, lactating (breast-feeding) women

#### 7] Study Assessments

Clinical assessment will be carried out at SGH.

Patient charts will be monitored closely for clinical symptoms and details will be recorded daily to record any adverse reactions. Fever, Radiographic evidence of pneumonia, Myalgia, Fatigue, Dyspnea, Tachycardia, Renal impairment (urine output), Septic shock, length of ICU admission, ventilator support, cardiac injury and any deterioration will be recorded.

Viral load will be determined by RT-PCR from nasopharyngeal swabs. The $C_T$ value at time of admission will be considered at 100%. From there the viral titer on Day 7 and day 15 will be estimated and calculated as % of day 0.

IFN-gamma, TNF-alpha and IL-6 will be determined by ELISA.

IgG and IgM will be estimated using ELISA for human samples.

Samples from patients will be kept frozen for future analyses of other parameters.

**Study Conduct**

Patient charts will be monitored closely for clinical symptoms and details will be recorded daily to record any adverse reactions. Fever, Radiographic evidence of pneumonia, Myalgia, Fatigue, Dyspnea, Tachycardia, Renal impairment (urine output), Septic shock, length of ICU admission, ventilator support, cardiac injury and any deterioration will be recorded.

| | | |
|---|---|---|
| 1. | Travel history | Yes |
| 2. | Medical history: | Yes (Attached) |
| 3. | Type of physical examination: | Breathlessness, Cough, |
| 4. | Symptom measurement: | Fever |
| 5. | Diagnostic testing: | Negative for HIV, HBV, |
| 6. | Chest X Ray | Yes |
| 7. | Blood or urine testing: | Yes |
| 8. | Tachycardia | Yes |
| 9. | Subject cohort assignment: | Yes |
| 10. | Adverse event review, etc. | Yes |
| 11. | Blood draw | Day 0, day 7 and 15 after BCG intervention |
| 12. | Day of ICU admission | Yes |
| 13. | Day of ventilator support and hours | Yes |
| 14. | Day of hospitalization | Yes |

Samples from patients will be kept frozen for future analyses of other parameters.

**Discontinued Subjects:**

***Describes the circumstances for Subject withdrawal, dropouts, or other reasons for discontinuation of Subjects. State how dropouts would be managed and if they would be replaced.***

Subject enrolled may decide to withdraw from the trial or primary attending physician make make the decision to violate protocol for best interest of patient. In such a case, we will partially analyze such data until date of withdrawal etc.

***Describe the method of handling of protocol waivers, if any. The person(s) who approves all such waivers should be identified and the criteria used for specific waivers should be provided.***

Primary attending physician along with Dean, BJ Medical College & SGM will decide protocol waivers, introduction of other drugs during the time period etc. In such a case, data will not be considered for analyses.

***Describes how protocol violations will be treated, including conditions where the study will be terminated for non-compliance with the protocol.***

If 3 subjects out of 30 subjects in experimental group continuously display serious adverse events leading to deterioration of patient and interventions other than concomitant drugs, the study will be terminated immediately.

#### 8] Study Treatment

**Experimental arm**

**DOSAGE**:

BCG will be administered in the dose of 0.1 ml, once during the entire period of the trail on day of admission.

**ROUTE OF ADMINISTRATION:** Intradermal Injection

**Control Arm**

Saline will be administered 0.1 ml, once during the entire period of the trail on day of admission.

**(b) Study drug supplies and administration:**

***A statement about who is going to provide the study medication and that the investigational drug formulation has been manufactured following all regulations. Details of the product stability, storage requirements and dispensing requirements should be provided.***

Haffkine Institute for Training, Research & Testing will provide Tubervac and Placebo (sodium chloride I.p.) Tubervac produced by Serum Institute of India has been approved by the Universal vaccination scheme for India. Tubervac should be stored in the dark between 2-8 degC. Diluent will not be frozen but will be stored cool (as declared by manufacturer). Reconstituted Tubervac lasts for 6 hours. We will use a fresh vial of Tubervac for each patient.

UnconstitutedTubervac24 months from the date of last satisfactory potency test if stored in a dark place at recommended temperature (Shelf life as declared by manufacturer).

Standard of care will be provided by BJ Medical College

***(c) Dose modification for study drug toxicity: Rules for changing the dose or stopping the study drug should be provided.***

Not applicable

**543 DrugsandCosmeticsRules,1945**

***(d) Possible drug interactions.***

Azithromycin reduces the effectiveness of BCG

Very minor drug interaction with Chloroquine

**(e) Concomitant therapy: The drugs that are permitted during the study and the conditions under which they may be used are detailed here.**

**Standard of care drugs.**

***Describe the drugs that a subject is not allowed to use during parts of or the entire study. If any washout periods for prohibited medications are needed prior to enrolment these should be described here.***

Subjects on

1. Immuno-suppressants
2. Systemic lupus (Belimumbab)
3. Anti-retroviral therapy (ART),
4. Chemotherapeutic (e.g. Cisplatin, Placitaxel) drugs
5. Radiation therapy
6. A combination of one or more of the above

Subjects with washout periods for any drugs (e.g. blood thinners, aspirin delayed response for ischemic heart etc) will also not be enrolled.

Subjects will not be given immuno-suppressants during the course of the study.

Subjects will not undergo blood transplant or plasma treatments during the course of the study.

***(f) Blinding procedures: A detailed description of the blinding procedure if the study employs a blind on the Investigator and/or the Subject***.

The proposed trial will be a randomized double blind trial.

BCG and Saline will be kept in identical looking bottles and coded separately. Neither attending physicians nor subjects will know their status.

Subjects falling within the inclusion criteria will be alternatively enrolled for placebo or test drug.

Subjects will be read out the informed consent form by concerned personnel.

***(g) Unblinding procedures: If the study is blinded, circumstances in which unblinding may be done and the mechanism to be used for unblinding should be given.***

Unblinding will be carried out in the vent of unforeseen eventualities to protect subjects in the event of medical or safety reasons, e.g. during an acute reaction. The unblinding information will be shared only on a need-to-know basis and 'break the blind' process for a single participant will be followed.

#### 9] Adverse Events

As already noted earlier, if 3 subjects in the experimental arm continuously show deterioration, the study will be terminated immediately.

TUBERVAC is contraindicated in hypogamma-globulinemia, congenital immunodeficiency, sarcoidosis, leukaemia, generalised malignancy, HIV infections or any other disorder in which natural immune response is altered, as also those on immunosuppressive therapy, corticosteroids, radiotherapy. Inchronic eczema or other dermatological disease, the vaccine can be given in a healthy area of the skin. Keloid and lupoid reactions may also occur at the site of injection and such children should not be revaccinated

*Skin lesions distinct from the vaccination site*. Tuberculosis infection can cause a number of cutaneous lesions (such as TB chancre, lupus vulgaris, scrofuloderma, papulonecrotictuberculidsetc). There are case reports of cutaneous lesions, distinct from the site of vaccination, thought to have occurred after BCG vaccination (Bellet et al., 2005). It is important to note that multiple cutaneous lesions may signal disseminated BCG disease usually in an immunocompromised host. There are case reports of lupus vulgaris, scrofuloderma following BCG vaccination.

*Lymphadenitis*. When severe, this includes nodes which become adherent to overlying skin with or without suppuration. Suppuration has been defined as "presence of fluctuation on palpation or pus on aspiration, the presence of a sinus, or large lymph node adherent to the skin with a caseous lesions on excision" (Lotte et al., 1984). If BCG is administered in the recommended site (deltoid) the ipsilateral axillary nodes are most likely to be affected but supra-clavicular or cervical nodes may also be involved (Hengster et al., 1992). The onset of suppuration may be variable with cases presenting from one week to 11 months following vaccination (de Souza et al., 1983). Lymphadenitis presenting within 2 months of vaccination and larger nodes (+ 1cm) may be less likely to resolve spontaneously (Caglayan et al., 1991). Suppurative lymphadenitis is now rare, especially when BCG inoculations are performed by well-trained staff, with a standardized freeze-dried vaccine and a clearly stated individual dose depending on the age of the vaccinated subjects.

*Osteitis and Osteomyelitis*. This is a rare and severe complication of BCG vaccination which has primarily been reported in Scandinavia and Eastern Europe and typically associated with changes in BCG vaccine strain. There was a report of an increase in osteitis to 35 per million in Czechoslovakia after a shift from the Prague to Russian strain BCG (Lotte, et al., 1988). Both Finland and Sweden reported increases in osteitis after 1971 when they shifted to a Gothenburg strain produced in Denmark. Sweden reported rates as high as 1 in 3,000 vaccine recipients, which declined rapidly when the national programme shifted to a Danish (Copenhagen, 1331) vaccine strain (Lotte et al., 1988). More recently reports of osteitis have become infrequent.

*Disseminated BCG disease or systemic BCG-itis.* This recognized but rare consequence of BCG vaccination traditionally has been seen in individuals with severe cellular immune deficiencies. The risk (fatal and non-fatal) is thought to be between 1.56 and 4.29 cases per million doses (Lotte et al., 1988). This is based on pre-HIV data. However, the exact incidence is debated because few centers are able to differentiate *Mycobacterium Bovis*BCG from other forms of Mycobacterium in patients presenting with disseminated disease. In a recent retrospective case series review of Mycobacteriunm tuberculosis complex 5% of cases were found to have the *M. Bovis*BCG strain (Hesseling et al., 2006). Additional data from studies in South Africa confirm the significantly high risk of disseminated BCG (dBCG) disease in HIV-positive infants, with rates approaching 1% (Hesseling et al., 2009).

As expected the cellular primary immunodeficiency predisposes to the condition. This includes severe combined immunodeficiency, chronic granulomatous disease, Di George syndrome and homozygous complete or partial interferon gamma receptor deficiency (Jouanguy et al., 1996; Jouanguy et al., 1997; Casanova et al., 1995).

In one series of 60 cases of BCG-itis the case fatality rate was approximately 50% although other smaller studies have documented a higher mortality rate (Lotte et al., 1988, Talbot et al., 1997). Early recognition and diagnosis is critical to management. In patients with primary immunodeficiency disorders the disease may be fatal without reconstitution of immunity through stem cell transplant.

*Immune reconstitution inflammatory syndrome (IRIS).* This has recently been identified as a BCG vaccine-related adverse event in immunocompromised individuals due to HIV started on antiretroviral therapy (ART) (DeSimone et al., 2000). It usually presents within 3 months of immune restoration and manifests as local abscesses or regional lymphadenitis usually *without* dissemination. No fatal cases have yet been documented. The etiology is unknown but postulated to be a deregulated inflammatory reaction directed against opportunistic pathogens including mycobacterial organisms following immune reconstitution.

*Other rare events.* A number of events have been reported as case reports or series. These include sarcoidosis, ocular lesions (conjunctivitis, choroiditis, optic neuritis), and erythema nodosum. Tuberculous meningitis (due to the BCG) has been described but is exceptionally rare (Tardieu et al., 1988)

#### 10] Ethical Considerations

***(a) Risk/benefit assessment:***

**Risks**

1) BCG infections have been reported in health care workers, primarily from exposures resulting from accidental needle sticks or skin lacerations during the preparation of BCG for administration.

2) Between 2004 and 2006, the incidence of TB in HIV-infected infants in Cape Town, South Africa was reported as 1596 per 100 000 compared to 66 per 100 000 in HIV-uninfected infants. This indicated that there is a substantiated higher risk of disseminated BCG disease developing in children infected with HIV who are vaccinated at birth and who later developed AIDS.

Most other risks in administration of BCG are due to poor immune status.

Both above groups of subjects are not part of the population as stated in the Inclusion criteria

**Benefits**

BCG is known to confer general immunity to patients, specifically through increase in Interferon -gamma. The kinetics of Interferon increase after BCG re-vaccination is debated and reports suggest that it varies from 7 days to 4 weeks.

Presently the average stay of a patient with mild COVID-19 in the hospital is around 15 days.

Should BCG re-vaccination cause an early spike in IFN-gamma, that would prove beneficial to the patient in controlling viremia, it would mean better outcomes and decrease duration of hospitalization and associated stress.

***(b) Ethics Committee review and communications.***

Approval Certificate Attached

***(c) Informed consent process.***

Attached

***(d) Statement of Subject confidentiality including ownership of data and coding procedures.***

Samples will be coded serially. Subjects will be allotted into the experimental arm and control arm by loading identical syringes with saline and BCG and blind choosing by treating physician. Confidentiality maintained by keeping records on an on-site computer.

#### 11] Study Monitoring and Supervision

***Case Record Form(CRF) completion requirements, including who gets which copies of the forms and any specifics required in filling out the forms CRF correction requirements, including who is authorized to make corrections on the CRF and how queries about study data are handled and how errors, if any, are to be corrected should be stated. Investigator study files, including what needs to be stored following study completion should be described.***

CRF completion responsibility: Site Investigators or delegated staff. Please find attached CRF attached.

#### 12] Investigational Product Management

***(a) Give Investigational product description and packaging (stating all Ingredients and the formulation of the investigational drug and any placebos used in the study)***

Participants will receive a single dose of BCG vaccine (Tubervac). The adult dose of BCG vaccine is 0.1 mL injected intradermally over the distal insertion of the deltoid muscle onto the humerus (approximately one third down the upper arm).

**COMPOSITION:** Live, attenuated BCG Vaccine (Bacillus Calmette-Guerin Strain) Each 1 ml contains between $1 \times 10^6$ and $33 \times 10^6$ Colony Forming Units (C.F.U.)

**Diluent:** Sodium Chloride Injection I.P.

***(b)The precise dosing required during the study.***

The adult dose of BCG vaccine is 0.1 mL injected intradermally

***(c) Method of packaging, labelling, and blinding of study substances.***

BCG injection will be prepared not in presence of the subject. Subject will be administered injection of BCG

***(d) Method of assigning treatments to Subjects and the Subject identification code***

Regarding subject identification, a study number will be assigned sequentially to included subjects.

***(e)Storage conditions for study substances.***

Tubervac should be stored in the dark between 2-8 degC. Diluent will not be frozen but will be stored cool (as declared by manufacturer).

Reconstituted Tubervac lasts for 6 hours. We will use a fresh vial of Tubervac for each patient.

UnconstitutedTubervac24 months from the date of last satisfactory potency test if stored in a dark place at recommended temperature (Shelf life as declared by manufacturer).

***(f) Investigational product accountability: Describe instructions for the receipt, storage, dispensation, and return of the investigational products to ensure a complete accounting of all investigational products received, dispensed, and returned/destroyed.***

Tubervac should be stored in the dark between 2-8 degC. Diluent will not be frozen but will be stored cool (as declared by manufacturer).

Reconstituted Tubervac lasts for 6 hours. We will use a fresh vial of Tubervac for each patient.

UnconstitutedTubervac24 months from the date of last satisfactory potency test if stored in a dark place at recommended temperature (Shelf life as declared by manufacturer).

***(g.) Describe policy and procedure for handling unused investigational products.***

If unused and un-reconstituted, BCG will be donated to the hospital for future immunizations of children. Reconstituted BCG is only stable for 6 hrs. So if unused, it will be discarded.

**13] Data Analysis**

***Provide details of the statistical approach to be followed including sample size, how the sample size was determined, including assumptions made in making this determination, efficacy endpoints (primary as well as secondary) and safety end points.***

***Statistical analysis: Give complete details of how the results will be analyzed and reported along with the description of statistical tests to be used to analyze the primary and secondary end points defined above.***

Results will be analyzed using Graph Pad Prism, 2 way Anova, t test, Mann Whitney test will be applied as applicable.

***Describe the level of significance, statistical tests to be used, and the methods used for missing data; method of evaluation of the data for treatment failures, non-compliance, and subject withdrawals; rationale and conditions for any interim analysis if planned.***

Level of significance will be as per parameter used. In case of RT-PCR data to evaluate viremia, samples will be run in triplicates,

For cytokines, ELISA will be used to evaluate IFN-gamma, TNF-alpha and IL-6 and each samples will be done atleast in duplicate

***Describe statistical considerations for Pharmacokinetic(PK) analysis, if applicable***

Not applicable

**14] Undertaking by the Investigator**

Attached

**15] Informed Consent Documents**

Attached

**16] Undertaking by the Sponsor/Sponsors representative/applicant to the licensing authority to provide medical management and compensation in case of clinical trial related injury or death for which subjects are entitled to compensation as required under rule 122DAB(6).**

We are working on an insurance policy/indemnity, see Invoice attached. Final certificate should be available in a day or two.

**17] Declaration regarding financial status of the applicant vis-à-vis medical management and compensation to be paid to the trial participants (in case of injury or death in clinical trial)**

Not applicable, since subjects are already and will receive free care for COVID-19 at BJ and Sassoon, sponsored by the Government.

**18] List of Investigators including site address (es).**

***Overall co-ordinator***

Dr. Sanjay Mukherjee, Hon. Prin Secretary, Medical Education & Drugs Department, 9th floor Mantralay, GT Hospital Campus, Fort, Mumbai 400 001.

Dr. Rajesh Deshmukh, Director, Haffkine Institute for Training, Research & testing, Acharya Donde Marg, Parel, Mumbai 400 012

***Scientific***

Dr. Usha Padmanabhan, Haffkine Institute for Training, Research & Testing, Acharya Donde Marg, Parel, Mumbai 400 012

Dr. Shashikant Vaidya, Haffkine Institute for Training, Research & Testing, Acharya Donde Marg, Parel, Mumbai 400 012

***Scientific***

Dr.Sonali Salvi, BJ Govt. Medical College & Sassoon General Hospital, Jayprakash Narayan Road, Near Pune Railway Station, Pune 411001

Dr. Rohidas Borse, BJ Govt. Medical College & Sassoon General Hospital, Jayprakash Narayan Road, Near Pune Railway Station, Pune 411001

Dr. Samir Joshi, BJ Govt. Medical College & Sassoon General Hospital, Jayprakash Narayan Road, Near Pune Railway Station, Pune 411001

Dr. Sanjay Gaikwad, BJ Govt. Medical College & Sassoon General Hospital, Jayprakash Narayan Road, Near Pune Railway Station, Pune 411001

Dr. Yogesh Gawali, BJ Govt. Medical College & Sassoon General Hospital, Jayprakash Narayan Road, Near Pune Railway Station, Pune 411001

Dr. Harish Tatia, BJ Govt. Medical College & Sassoon General Hospital, Jayprakash Narayan Road, Near Pune Railway Station, Pune 411001

(a) Trial site details

***Trial Site Address:***

BJ Govt. Medical College & Sassoon General Hospital, Jayprakash Narayan Road, Near Pune Railway Station, Pune 411001

***Whether Trial Site is equipped with super specialty or multi-specialty facilities and emergency facilities with Institutional ethics committee.***

Yes

**19] Ethics Committee approvals, if available:-(Institutional Ethics Committee should be in same area where the site is located).**

Yes. Approval certificate of Ethics committee of B. J. Govt. Medical College, Pune & SGH

**20] As per the protocol, whether the subjects will receive the standard care. (Give declaration)**

Yes Subjects will receive free standard care for COVID-19 at B. J. Govt. Medical College, Pune & SGH.

**21] Details of Contract entered by Sponsor with Investigating Institutions**

MoU has been executed Attached.
